# Sociodemographic inequalities of suicide: a population-based cohort study of adults in England and Wales 2011-2021

**DOI:** 10.1101/2023.04.05.23288190

**Authors:** Isobel L. Ward, Katie Finning, Daniel Ayoubkhani, Katie Hendry, Louis Appleby, Emma Sharland, Vahé Nafilyan

**Affiliations:** Office for National Statistics, UK; Division of Psychology and Mental Health, University of Manchester, UK

## Abstract

**Background:** Risk of suicide is complex and often a result of multiple interacting factors. It is vital research identifies predictors of suicide to provide a strong evidence base for targeted interventions.

**Methods:** Using linked Census and population level mortality data we estimated rates of suicide across different groups in England and Wales and examine which factors are independently associated with the risk of suicide.

**Findings:** The highest rates of suicide were amongst those who reported an impairment affecting their day-to-day activities, those who were long term unemployed or never had worked, or those who were single or separated. Rates of suicide were highest in the White and Mixed/multiple ethnic groups compared to other ethnicities, and in people who reported a religious affiliation compared with those who had no religion. Comparison of minimally adjusted models (predictor, sex and age) with fully-adjusted models (sex, age, ethnicity, region, partnership status, religious affiliation, day-to-day impairments, armed forces membership and socioeconomic status) identified key predictors which remain important risk factors after accounting for other characteristics; day-to-day impairments were still found to increase the incidence of suicide relative to those whose activities were not impaired after adjusting for employment status. Overall, rates of suicide were higher in men compared to females across all ages, with the highest rates in 40-to-50-year-olds.

**Interpretation:** The findings of this work provide novel population level insights into the risk of suicide by sociodemographic characteristics. Understanding the interaction between key risk factors for suicide has important implications for national suicide prevention strategies.

**Funding:** This study received no specific funding.

**Research in context:** *Evidence before this study:* Previous studies have identified key risk factors for suicide; being male and being aged 40 to 50 years of age have the highest rates of suicide. Suicide is a major public health concern, with prevention strategies imperative to minimising events.

*Added value of this study:* For the first time we make population level estimates of suicide rates in England and Wales using death registration data linked to 2011 Census. Furthermore, we calculate incidence rate ratios for fully adjusted models which provide novel insights into the interplay between different risk factors. For instance, we see that people who report having day-to-day impairments risk of suicide is 2- to 3-times higher for men and women respectively compared to people who do not report day-to-day impairments, after adjusting for other characteristics such as socioeconomic status which are likely associated with impairments.

*Implications of this study:* Understanding the groups most at risk of suicide is imperative for national suicide prevention strategies. This work provides novel population level insights into the risk of suicide by sociodemographic characteristics.

## Introduction

Suicide is a major public health concern and a leading cause of death globally, responsible for more than 700,000 deaths each year [1]. In England, there are over 5000 suicides annually [2] and in 2012 a National Suicide Prevention strategy was published for the first time, with key areas of action including reducing the risk of suicide among high-risk groups and supporting research, data collection and monitoring of suicide in England [3]. The strategy, updated in 2017, emphasised the importance that quality data and linkage of sources are utilised to provide the best estimates of at-risk groups who should be targeted for intervention. In 2021 the World Health Organization (WHO) called for improved monitoring of suicide to support development and implementation of effective suicide prevention strategies [4]. The development of suicide risk is complex and often includes a combination of biological, psychological, clinical, social, and environmental risk factors [5]. Previous research has highlighted that sociodemographic risk factors for suicide include being male, being middle- and old-age, and belonging to a low socioeconomic group [2], [6], [7]. However, few studies have investigated sociodemographic risk factors for suicide in England, and none with population level data which can be used to account for confounding differences between at risk groups. In the current work we use a novel linkage of 2011 Census and population level mortality data and modelling to assess which risk factors are important predictors of suicide.

## Methods

### Study Data and Population

We used Census 2011 for England and Wales and Mortality data linked by NHS number. To obtain NHS numbers, the 2011 census was linked to the 2011-13 NHS Patient Registers. A total of 50,189,388 individuals who were valid Census respondents and could be linked were included in our sample. Of these, 50,189,220 were either alive at the end of study (31^st^ December 2021) or died between 28^th^ March 2011 and end of study. Our final sample comprised of 35,136,916 people were between the ages 18 and 74 years on census day [Supplementary table 1]. We included adults up to age 74 years on 2011 Census day as National Statistics Socio-economic Classification (NS-SEC) was constructed to differentiate positions within labour markets in people aged 16 to 74.

**Table 1.** All covariates and groupings. All variables are defined from 2011 Census data.

| Variable | Levels |
| --- | --- |
| Sex | Male, Female |
| Age | Continuous spline in years calculated on Census Day |
| Ethnicity<sup>1</sup> | Arab,<br>Caribbean, African, Black British and other Black,<br>Chinese,<br>Indian,<br>Mixed/multiple ethnic groups,<br>Other ethnic group<br>Pakistani and other Asian/Asian British,<br>White |
| Partnership status | In Partnership (Married or in a registered same-sex civil partnership),<br>Partner deceased (Widowed or surviving partner from a same-sex civil partnership),<br>Separated (divorced, separated but still legally in a same-sex civil partnership or formally in a same-sex civil partnership which is now legally dissolved),<br>Single (never been legally married or never registered for a same-sex civil partnership) |
| Day-to-day impairments | Day to day activities limited a lot/a little,<br>Day to day activities not limited, Unknown |
| Religion | Buddhist, Christian, Hindu, Jewish, Muslim, No religion, Not stated, Other religion, Sikh |
| Region | East of England, North East, North Midlands, North West, South East, South West, Wales, West Midlands, Yorkshire and the Humber, London, Unknown |
| National Statistics Socio-economic Classification (NS-SEC) <sup>2</sup> | Higher managerial, administrative and professional occupations for large employers and higher managerial and administrative occupations (Class 1.1), higher professional occupations (Class 1.2), lower managerial, administrative and professional occupations (Class 2), intermediate occupations (Class 3), small employers and own account owners (Class 4), lower supervisory and technical occupations (Class 5), semi-routine occupations (Class 6), routine occupations (Class 7) and never worked and long term unemployed (Class 8). |

*Footnotes:* 1. Groupings of ethnic categories were based on GSS harmonised standard [1]; 2. National Statistics Socio-economic status analytic classes [2].
[1] Ethnicity Harmonised Standard, GSS Harmonisation Team, August 2011.
https://analysisfunction.civilservice.gov.uk/policy-store/ethnicity-harmonised-standard/#presentation-united-kingdom
[2] SOC2010 volume 3: the National Statistics Socio-economic classification (NS-SEC rebased on SOC2010), Office for National Statistics.
https://www.ons.gov.uk/methodology/classificationsandstandards/standardoccupationalclassifications/soc2010/soc2010volume3thenationalstatisticssocioeconomicclassificationnssecbasedonsoc2010#analytic-classes-and-operational-categories

In England and Wales, when someone dies unexpectedly, a coroner investigates to establish the cause of death through an inquest, resulting in a registration delay. For deaths caused by suicide, this generally means that around half of the deaths registered each year will have occurred in the previous year or earlier. To reduce the potential bias by this registration delay we used deaths occurring up to 31st December 2021 and registered by 31^st^ December 2022.

### Exposures

We investigated a range of potential sociodemographic factors likely to be associated with the risk of suicide, selected based on the existing literature [5], [8], and data availability. Exposure variables explored in this analysis were sex, age (as a natural spline with boundary knots at the 1^st^ and 99^th^ percentile and four interior knots), ethnicity, marital status, day-to-day impairments, religion, region, and NS-SEC [Table 1]. All exposures were self-reported from 2011 Census and age was calculated on census day.

### Outcome

Our primary outcome was death due to suicide at any time during the study follow-up period (28^th^ March 2011 to 31 December 2021). Suicide was defined as deaths from intentional self-harm for persons aged 10 years and over, and deaths caused by injury or poisoning where the intent was undetermined for those aged 15 years and over [2]. International Classification of Diseases, Tenth Revision (ICD-10) codes corresponding to intentional self-harm (X60-X84) and injury/poisoning of undetermined intent (Y10-Y34) were used.

### Statistical analysis

Descriptive statistics of population characteristics were presented stratified by all outcomes (alive, death by suicide, death due to all other causes). To model the association between the risk of suicide and demographic and socioeconomic characteristics, we fitted generalised linear models with a Poisson link function, with death by suicide being the outcome of interest. The natural logarithm of exposure time was included in the models as an offset term to account for the different time-at-risk periods between individuals. First, for each exposure, to estimate the difference in the rate of suicide, we fitted models adjusted for age and sex, with sex being interacted with age and the exposure of interest. The number of interior knots on the age spline was determined using the Bayesian Information Criterion (BIC). To estimate rates of suicide per 100,000 people for each level of the exposure, by sex for the average age, we calculated marginal means using the model with the lowest BIC. Estimated rates of suicide are reported per 100,000 people over the 10-year study period. Second, to understand the different risk across the life course, we tested for an interaction between the factor of interest and age. We used the BIC to assess model fit between the models.

Finally, to assess how each factor is independently associated with the risk of suicide, we fitted fully adjusted models. For each exposure, we compared the incident rate ratios (IRRs) from the fully adjusted model to those from the age and sex adjusted model. For both minimally and fully adjusted models, sex was interacted with the exposure of interest, and an age-sex interaction was included. IRRs were calculated separately for each exposure for males and females.

Follow-up time was calculated as the time from 27^th^ March 2011 (Census day) to date of death or end of study (31^st^ December 2021), whichever was earlier. All statistical analyses were performed using R version 3.5.1.

## Results

### Estimated rates of suicide per 100,000 people

Our study population consisted of 35,136,917 people; there were 35,928 suicides in our study period, with 73.9% of these suicides occurring in men [Supplementary Table 2]. Estimated rates of suicide were higher in men (19.8 per 100,000 people, 95% confidence interval (CI):19.3-20.2) compared with women (6.5 per 100,000 people, 95%CI:6.2-6.7), with the highest in males aged 40-to-50-years [Figure 1]. In women, the rates of suicide were highest in those aged 45-to-50-years but remained lower than men across all age groups.

**Figure 1.**
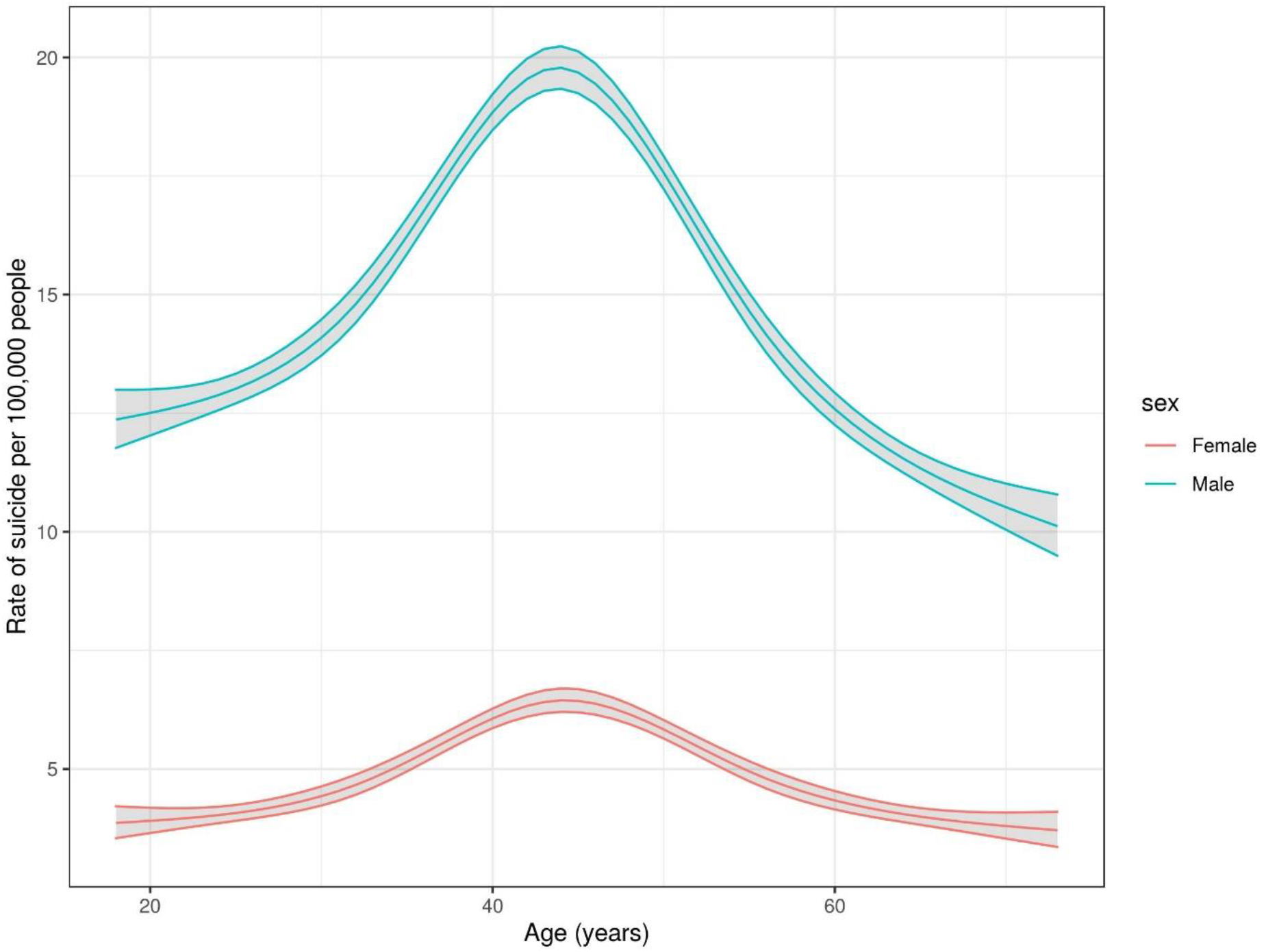
Estimated rates of suicide per 100,000 people by age and sex. Estimated rates of suicide per 100,000 people by age and sex from a Poisson model. Age was interacted with sex. Age was included as a natural spline with boundary knots at the 1^st^ and 99^th^ percentiles and four internal knots.

For all exposures, except day-to-day impairments, the model without interaction between the exposure and age had a lower BIC than the model with the interaction. For day-to-day impairments, the fully interacted model (sex, age and day-to-day impairments) was the best fit. The estimated rates of suicide per 100,000 people for each exposure were estimated from the best model fit. Rates have been calculated for each level of the exposure, by sex for several ages (20, 30, 40, 50, 60 and 70 years of age) when the size of the denominator population is greater than 30. For the armed forces variable rates were only estimated up to age 50 years due to the underlying population distribution. In addition, for each model the rate was estimated by sex for the average age in our study population (45-years-of-age).

For NS-SEC, the highest rates of suicide were seen in people who had never worked and or were long-term unemployed (men: 37.1 per 100,000 people, 95%CI:35.1-39.3, women: 12.0 per 100,000 people, 95% CI:11.0-13.1) [Table 2, Figure 2]. Those classified as having higher managerial, administrative, and professional occupations (Class 1.1) had the lowest rates (men: 12.6 per 100,000 people, 95% CI:11.6-13.7, women: 4.6 per 100,000 people, 95%CI:4.0-5.2). For estimated rates of suicide by age for each characteristic see Supplementary Table 3.

**Figure 2.**
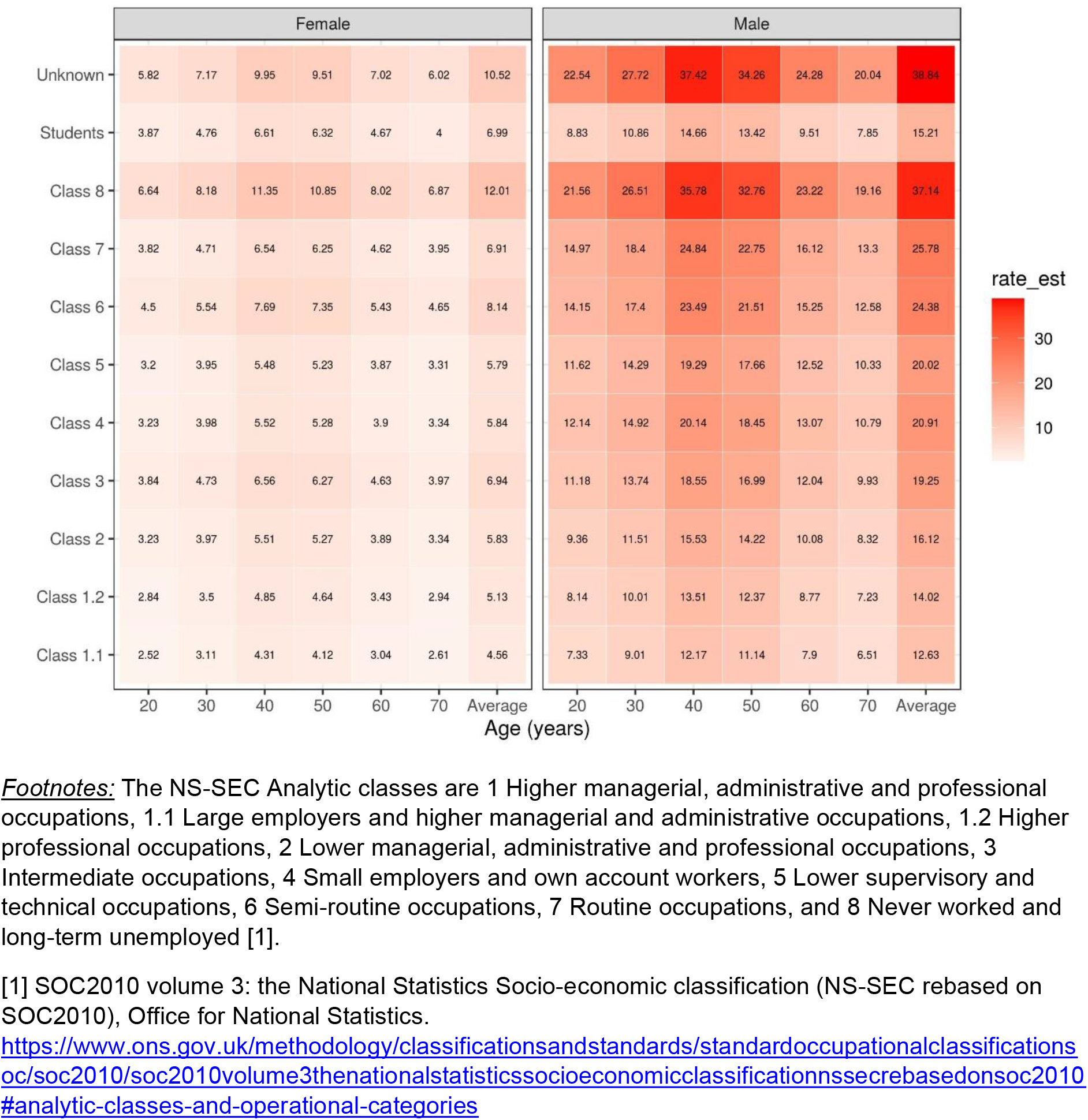
Estimated rates of suicide per 100,000 people by socio-economic status. Estimated rates of suicide per 100,000 people by NS-SEC from a Poisson model with NS-SEC and age both interacted with sex.

**Table 2.**
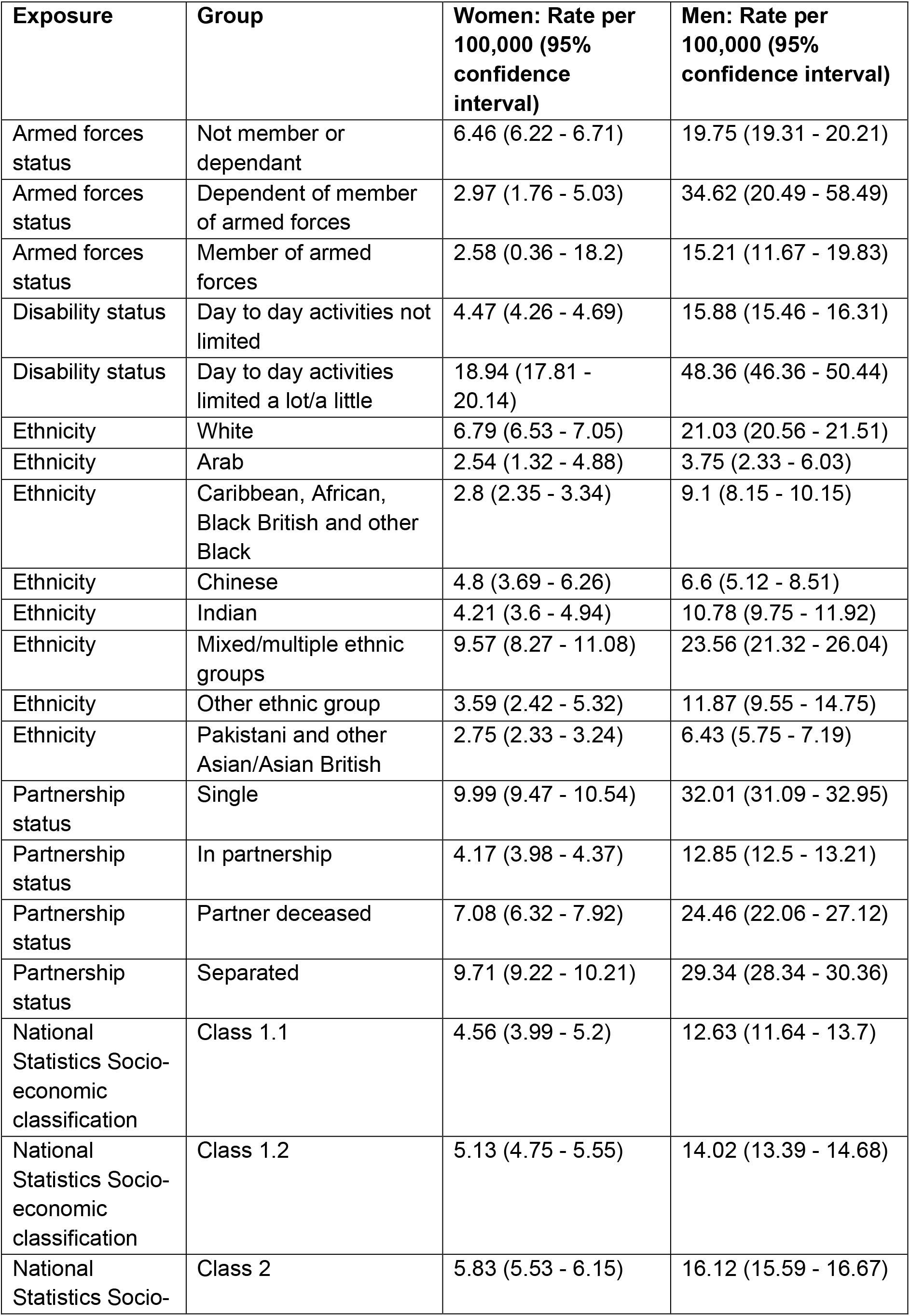
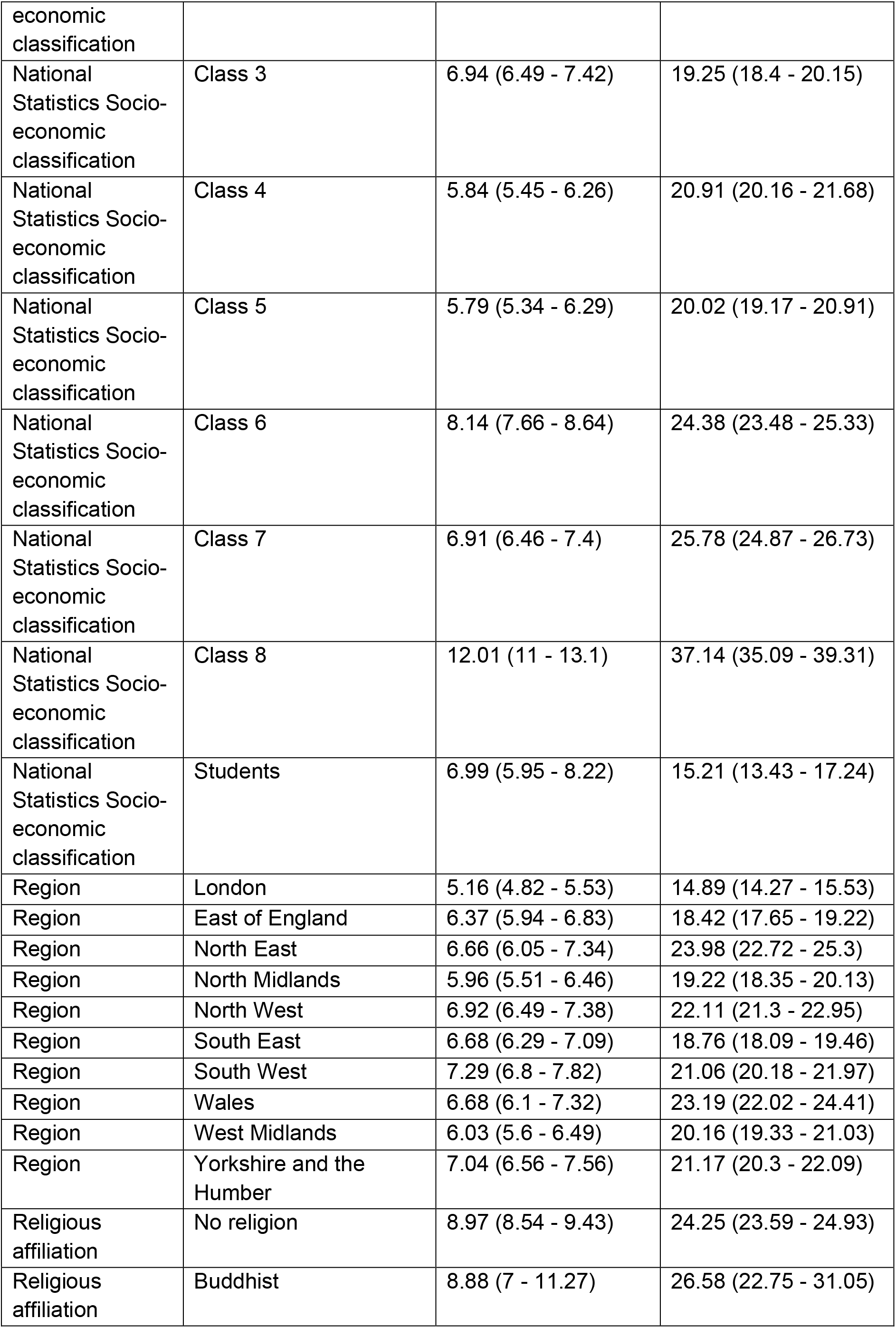
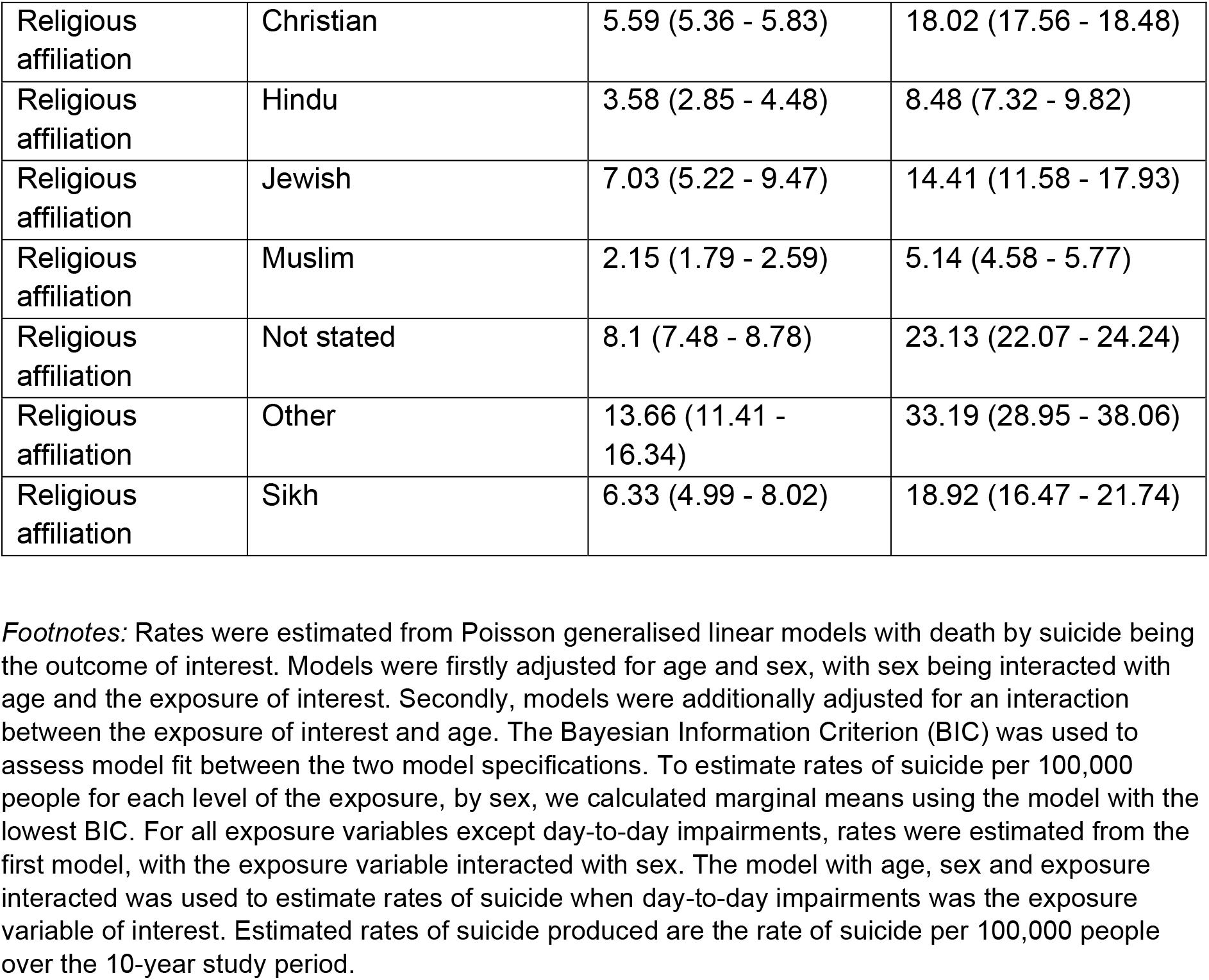
Estimated rates of suicide by sociodemographic characteristics for average age.

Estimated rates of suicide were highest in the White (men: 21.0 per 100,000 people, 95%CI:20.6-21.5, women: 6.8 per 100,000 people, 95%CI:6.5-7.1) and Mixed/Multiple ethnic groups (men: 23.56 per 100,000 people, 95%CI:21.3-26.0, women: 9.6 per 100,000 people, 95%CI:8.3-11.1). Estimated rates of suicide were lowest for the Arab group (men: 3.8 per 100,000 people, 95%CI:2.3-6.0, women: 2.5 per 100,000 people, 95%CI:1.3-4.9) [Table 2].

For religion, the lowest rates of suicide were in the Muslim group (men: 5.1 per 100,000 people, 95%CI:4.6-5.8, women: 2.2 per 100,000 people, 95%CI:1.8-2.6) (Figure 3). The rates of suicide were highest in the Buddhist group (men: 26.6 per 100,000 people, 95%CI:22.8-31.1, women: 8.9 per 100,000 people, 95%CI:7.0-11.3) and religions classified as “Other” (men: 33.2 per 100,000 people, 95%CI:29.0-38.1, women: 13.7 per 100,000 people, 95%CI:11.4-16.3) [Table 2]. For men and females, the rates of suicide were lower across the Muslim, Hindu, Jewish, Christian and Sikh groups compared to the group who do not have religious beliefs.

**Figure 3.**
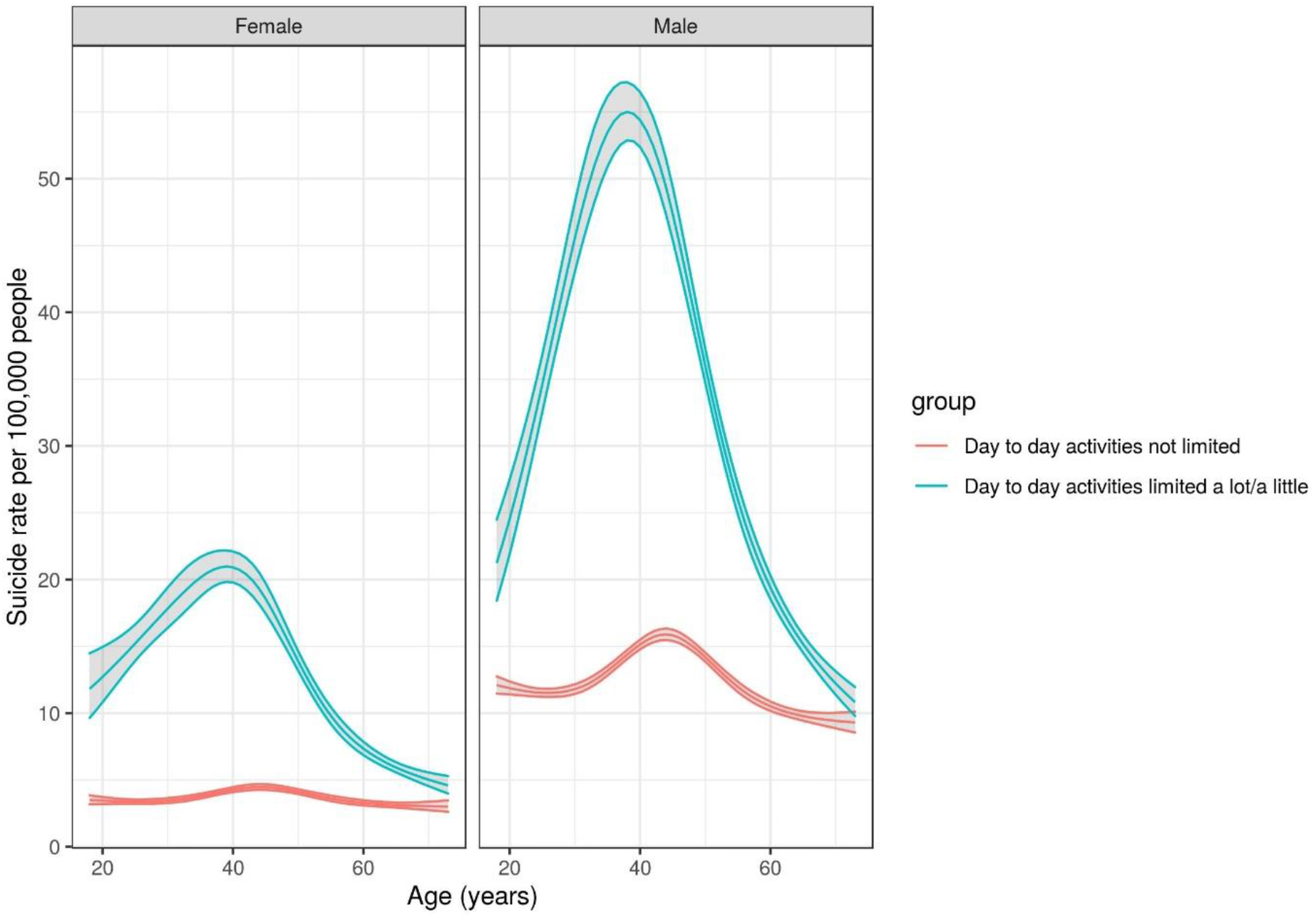
Rates of suicide per 100,000 people by day-to-day impairment. Estimated rates of suicide per 100,000 people by Day-to-day impairments forces status from a Poisson model with age, sex and Day-to-day impairments all interacted.

For marital status, people who described themselves as in a partnership, which is either married or in a registered same-sex civil partnership, had the lowest rates of suicide (men: 12.9 per 100,000 people, 95%CI:12.5-13.2, women 4.2 per 100,000 people, 95%CI:4.0-4.4) (Figure 4). This was when compared with people who described themselves as single, separated or partner deceased [Table 2].

Rates of suicide were lowest in London (men: 14.9 per 100,000 people, 95%CI:14.3-15.5, women: 5.2 per 100,000 people, 95%CI:4.8-5.5) and highest in the North East for men (24.0 per 100,000 people, 95%CI:22.7-25.3) and in the South West for women (7.3 per 100,000 people, 95%CI:6.8-7.8) [Table 2]. At the time of the 2011 Census, rates of suicide were lower among serving members of the armed forces, than non-members for both men and women [Table 2]. The rate was lower for women who were dependants of a serving member of the armed forces (3.0 per 100,000 people, 95%CI:1.8-5.0) compared with those who were not members or dependants of a member (6.5 per 100,000 people, 95%CI:6.2-6.7). Conversely, for men the opposite was found, with dependants of serving members of the armed forces having the highest rate compared with those who were current members or not members.

People who had day-to-day impairments had much higher rates of suicide (men: 48.4 per 100,000 people, 95%CI:48.4-50.4, women: 18.9 per 100,000 people, 95%CI:17.8-20.1) compared with non-disabled people (men: 15.9 per 100,00 people, 95%CI:15.5-16.3, women: 4.5 per 100,000 people, 95%CI:4.3-4.7) [Table 2, Figure 3]. We found the model which fully interacted age, sex and day-to-day impairments was the best fitting model; differences between those (both men and females) who reported day-to-day impairments were greater in younger individuals, with the difference in rates between groups being less visible in older individuals [Figure 3].

The groups with the highest rates of suicide in men were those in middle age who had day-to-day impairments, were unemployed, single, separated or part of a non-main religious group. Men aged 30-to-50-years with day-to-day impairments had the highest rate of suicide (Rates: 54.4 to 36.0 per 100,000 persons). The next most vulnerable group were men ages 40-to-50-years who were long term unemployed or never worked (Rates: 32.8 to 35.8 per 100,000 persons). For women, the rates of suicide were highest amongst disabled women aged 30 to 50 (Rates: 12.7 to 20.9 per 100,000 persons). The next highest risk group was females aged 40-to-50-years who identified with a non-main religious group (Rates: 12.6 to 12.7 per 100,000 persons).

### Independent association of the different sociodemographic characteristics

Comparison of IRRs between a fully adjusted model (age, sex, ethnicity, marital status, day-to-day impairments, region, NS-SEC, religion, and member of the armed forces status) and individual minimally adjusted models (each exposure plus age and sex) allowed us to identify independent associations been risk factors and suicide. Comparison of IRRs found that the values were generally closer to 1 in the fully adjusted model. For instance, the rate of suicide in the Mixed/multiple ethnic females was 1.4 times higher (IRR=1.4, 95%CI=1.1;1.6) compared to white females in the minimally adjusted model, however when accounting for other characteristics in the fully adjusted model the rate remained elevated, but the difference was closer to the reference group (IRR=1.3, 95%CI=1.1;1.5) [Table 3]. Interestingly, the IRRs for day-to-day activities being limited a lot compared to not limited (Men IRR=2.6, 95%CI=2.5;2.7, Women IRR=3.5, 95%CI=3.3-3.6), which is a marker of day-to-day impairments, remained elevated in the fully adjusted model (Men IRR=2.1, 95%CI=2.1;2.2, Women IRR=3.1, 95%CI=3.3;3.6 [Table 3].

**Table 3.** Adjusted incidence rate ratios for suicide by sociodemographic characteristic.

| **Group** | **Exposure** | **Women:**<br>**Minimally**<br>**adjusted**<br>**model**<br>**[Incidence**<br>**rate ratio**<br>**(95%**<br>**confidence**<br>**interval)]** | **Women:**<br>**Fully**<br>**adjusted**<br>**model**<br>**[Incidence**<br>**rate ratio**<br>**(95%**<br>**confidence**<br>**interval)]** | **Men:**<br>**Minimally**<br>**adjusted**<br>**model**<br>**[Incidence**<br>**rate ratio**<br>**(95%**<br>**confidence**<br>**interval)]** | **Men: Fully**<br>**adjusted**<br>**model**<br>**[Incidence**<br>**rate ratio**<br>**(95%**<br>**confidence**<br>**interval)]** |
| --- | --- | --- | --- | --- | --- |
| Arab | Ethnicity | 0.37 (0.19 - 0.72) | 0.69 (0.35 - 1.34) | 0.18 (0.11 - 0.29) | 0.36 (0.22 - 0.59) |
| Caribbean, African, Black British and other Black | Ethnicity | 0.41 (0.35 - 0.49) | 0.46 (0.38 - 0.55) | 0.43 (0.39 - 0.48) | 0.5 (0.45 - 0.56) |
| Chinese | Ethnicity | 0.71 (0.54 - 0.92) | 0.74 (0.56 - 0.96) | 0.31 (0.24 - 0.4) | 0.34 (0.26 - 0.44) |
| Indian | Ethnicity | 0.62 (0.53 - 0.73) | 0.73 (0.57 - 0.95) | 0.51 (0.46 - 0.57) | 0.64 (0.54 - 0.75) |
| Mixed/multiple ethnic groups | Ethnicity | 1.41 (1.22 - 1.63) | 1.28 (1.11 - 1.48) | 1.12 (1.01 - 1.24) | 1.08 (0.98 - 1.2) |
| Other ethnic group | Ethnicity | 0.53 (0.36 - 0.78) | 0.62 (0.41 - 0.92) | 0.56 (0.45 - 0.7) | 0.7 (0.56 - 0.87) |
| Pakistani and other Asian/Asian British | Ethnicity | 0.4 (0.34 - 0.48) | 0.66 (0.54 - 0.81) | 0.31 (0.27 - 0.34) | 0.57 (0.5 - 0.66) |
| In partnership | Partnership status | 0.42 (0.39 - 0.44) | 0.52 (0.49 - 0.55) | 0.4 (0.39 - 0.41) | 0.51 (0.5 - 0.53) |
| Partner deceased | Partnership status | 0.71 (0.63 - 0.8) | 0.79 (0.7 - 0.89) | 0.76 (0.69 - 0.85) | 0.84 (0.76 - 0.93) |
| Separated | Partnership status | 0.97 (0.91 - 1.03) | 1.01 (0.95 - 1.07) | 0.92 (0.88 - 0.95) | 0.98 (0.95 - 1.02) |
| Day to day activities limited a lot/a little | Day-to-day impairments | 3.49 (3.33 - 3.64) | 3.14 (3 - 3.28) | 2.58 (2.5 - 2.65) | 2.12 (2.06 - 2.18) |
| East of England | Region | 1.23 (1.13 - 1.35) | 1.16 (1.06 - 1.26) | 1.24 (1.17 - 1.31) | 1.09 (1.03 - 1.15) |
| North East | Region | 1.29 (1.16 - 1.44) | 1.1 (0.99 - 1.23) | 1.61 (1.51 - 1.71) | 1.29 (1.21 - 1.37) |
| North Midlands | Region | 1.16 (1.05 - 1.27) | 1.05 (0.95 - 1.16) | 1.29 (1.22 - 1.37) | 1.1 (1.04 - 1.16) |
| North West | Region | 1.34 (1.24 - 1.45) | 1.19 (1.09 - 1.29) | 1.49 (1.41 - 1.56) | 1.24 (1.18 - 1.3) |
| South East | Region | 1.29 (1.2 - 1.4) | 1.24 (1.14 - 1.34) | 1.26 (1.2 - 1.32) | 1.13 (1.08 - 1.19) |
| South West | Region | 1.41 (1.3 - 1.54) | 1.27 (1.16 - 1.39) | 1.41 (1.34 - 1.49) | 1.19 (1.12 - 1.25) |
| Wales | Region | 1.29 (1.17 - 1.44) | 1.05 (0.94 - 1.16) | 1.56 (1.47 - 1.66) | 1.21 (1.14 - 1.29) |
| West Midlands | Region | 1.17 (1.07 - 1.28) | 1.09 (0.99 - 1.19) | 1.35 (1.28 - 1.43) | 1.18 (1.12 - 1.25) |
| Yorkshire and the Humber | Region | 1.36 (1.25 - 1.49) | 1.24 (1.13 - 1.35) | 1.42 (1.35 - 1.5) | 1.2 (1.14 - 1.27) |
| Class 1.2 | NS-SEC | 1.13 (0.97 - 1.3) | 1.11 (0.96 - 1.29) | 1.11 (1.01 - 1.22) | 1.11 (1.01 - 1.21) |
| Class 2 | NS-SEC | 1.28 (1.12 - 1.47) | 1.12 (0.98 - 1.29) | 1.28 (1.17 - 1.39) | 1.19 (1.09 - 1.29) |
| Class 3 | NS-SEC | 1.52 (1.32 - 1.75) | 1.18 (1.02 - 1.36) | 1.52 (1.39 - 1.67) | 1.32 (1.21 - 1.44) |
| Class 4 | NS-SEC | 1.28 (1.11 - 1.48) | 1.18 (1.03 - 1.37) | 1.66 (1.52 - 1.8) | 1.51 (1.38 - 1.64) |
| Class 5 | NS-SEC | 1.27 (1.09 - 1.48) | 1.08 (0.93 - 1.25) | 1.59 (1.45 - 1.73) | 1.36 (1.24 - 1.48) |
| Class 6 | NS-SEC | 1.78 (1.55 - 2.05) | 1.29 (1.12 - 1.48) | 1.93 (1.77 - 2.1) | 1.57 (1.44 - 1.71) |
| Class 7 | NS-SEC | 1.52 (1.32 - 1.75) | 1.12 (0.97 - 1.29) | 2.04 (1.87 - 2.22) | 1.59 (1.46 - 1.73) |
| Class 8 | NS-SEC | 2.63 (2.26 - 3.07) | 1.49 (1.27 - 1.74) | 2.94 (2.67 - 3.24) | 1.88 (1.71 - 2.08) |
| Students | NS-SEC | 1.53 (1.25 - 1.88) | 1.18 (0.96 - 1.45) | 1.2 (1.04 - 1.4) | 1.13 (0.98 - 1.31) |
| Buddhist | Religion | 0.99 (0.78 - 1.26) | 1.31 (1.02 - 1.68) | 1.1 (0.94 - 1.28) | 1.39 (1.18 - 1.62) |
| Christian | Religion | 0.62 (0.59 - 0.65) | 0.69 (0.66 - 0.72) | 0.74 (0.72 - 0.76) | 0.78 (0.76 - 0.8) |
| Hindu | Religion | 0.4 (0.32 - 0.5) | 0.73 (0.53 - 1) | 0.35 (0.3 - 0.4) | 0.75 (0.61 - 0.92) |
| Jewish | Religion | 0.78 (0.58 - 1.06) | 1 (0.74 - 1.35) | 0.59 (0.48 - 0.74) | 0.77 (0.62 - 0.96) |
| Muslim | Religion | 0.24 (0.2 - 0.29) | 0.36 (0.28 - 0.45) | 0.21 (0.19 - 0.24) | 0.39 (0.34 - 0.45) |
| Not stated | Religion | 0.9 (0.83 - 0.98) | 0.93 (0.85 - 1.01) | 0.95 (0.91 - 1) | 0.95 (0.9 - 0.99) |
| Other | Religion | 1.52 (1.27 - 1.82) | 1.26 (1.05 - 1.51) | 1.37 (1.19 - 1.57) | 1.28 (1.11 - 1.47) |
| Sikh | Religion | 0.71 (0.56 - 0.89) | 1.16 (0.84 - 1.6) | 0.78 (0.68 - 0.9) | 1.45 (1.2 - 1.76) |
| Dependent of member of armed forces | Armed forces status | 0.46 (0.27 - 0.78) | 0.65 (0.38 - 1.1) | 1.75 (1.04 - 2.96) | 2.12 (1.25 - 3.58) |
| Member of armed forces | Armed forces status | 0.4 (0.06 - 2.82) | 0.43 (0.06 - 2.97) | 0.77 (0.59 - 1) | 0.88 (0.67 - 1.14) |

*Footnotes:* Incidence rate ratios for adults were estimated from Poisson generalised linear models with death by suicide being the outcome of interest. Minimally adjusted models were adjusted for age and sex, with sex interacted with age and the exposure variable. Fully adjusted models were adjusted for sex, age, ethnicity, partnership status, day-to-day impairments status, religious affiliation, region, National Statistics Socio-economic Classification (NS-SEC) and armed forces membership. The fully adjusted models also included interactions between age and sex, and sex and the exposure variable of interest. The incidence rate ratio for the reference group is always equal to 1, with the reference group for each variable as listed: White (ethnicity); single (partnership status); day-to-day activities not
limited (day-to-day impairments); no religion (religious affiliation); London (region); class 1.1 (NS-SEC); not member or dependent (armed forces membership).

The rate of suicide was highest for people in NS-SEC Class 8 who are long term unemployed (Men IRR=2.9, 95%CI=2.7;3.2, Women IRR=2.6, 95%CI=2.2;3.1) [Table 3]. We see a gradual increase in rates moving from analytical Class 1.1 to Class 8 highlighting that socio-economic status is an important indicator for magnitude of risk. When accounting for other differences the risk is significantly reduced, due to adjusting for other factors such as day-to-day impairments which are likely confounding (Men IRR=1.9, 95%CI=1.7;2.1, Women IRR=1.5, 95%CI=1.3;1.7) [Table 3]. After accounting for other factors, being Buddhist was associated with an elevated risk of suicide compared to having no religion (Men IRR=1.4, 95%CI=1.2;1.6, Women IRR=1.3, 95%CI=1.0;1.7) [Table 3]. For regional differences, after accounting for other factors such as ethnicity and socioeconomic status which are likely key drivers of population variation, disparities between regions were reduced but still remained with all regions having higher rates than London [Table 3].

## Discussion

This study explored the sociodemographic factors associated with the risk of suicide among 35 million adults between 2011 and 2021. To our knowledge, this is the first study to investigate sociodemographic factors associated with the risk of death by suicide in England and Wales using population level data. We found that several sociodemographic characteristics were strongly associated with the risk of suicide; being male, or being 40-to-50-years-of-age were found to have the highest risk of suicide. Reporting day-to-day impairments and never having worked or being long term unemployed were also found to be associated with an increased risk of suicide. Those belonging to the White or Mixed/Multiple ethnic groups had the highest rates compared to other ethnic categories.

We find the rate of suicide are 2.5-3.5 times greater in men and women reporting day-to-day impairments than those who do not. The relative rates are higher in women reporting day-to-day impairments versus those without compared to men, suggesting this group should be a target for intervention. Overall, we find that the relative risk of suicide remains significantly elevated in people who have day-to-day impairments even after adjusting for other factors, indicating that the risk is independently associated with impairments. Day-to-day impairments is likely capturing factors associated with multimorbidity, mental health and old age. The census question asked respondents if their day-to-day activities were limited because of a health problem or disability which has lasted or is expected to last more than 12 months. Our findings show people whose day-to-day activities are limited either a lot or a little, relative to those who are not limited, had higher rates of suicide. Some research has found that severe health conditions are associated with a higher risk of suicide [10], [11].

We find never having worked or being long term unemployed was associated with the highest rates of suicide in both men and women. Socioeconomic position has been found to be an important risk factor for suicide[14]. In our minimally adjusted model, the IRRs were 2.5-3 times higher for women and men who were long term unemployed or never worked compared to those who were in higher managerial, administrative, and professional occupations. When adjusting for other characteristics we see the relative risk is 1.5-1.8 times higher, suggesting that accounting for factors such as day-to-day impairments could be capturing some of this variability.

It is an established finding that suicides are lower in serving members of the armed forces compared to members of the general population [15].It may be due to the “healthy worker effect”; people in employment generally, and especially in physically demanding jobs like the armed forces, are less likely than the general population to be ill or disabled. In the England, recent work has shown that veterans are at no greater risk of suicide than the general population [16]. Our sample will likely capture individuals who since the time of 2011 Census are now veterans, as well as those who remained serving members at the end of study.

We find the rates are highest in men aged 40 to 50 years of age. Middle-aged and older men have been shown to have the highest rates of suicide [2], [7]. In June 2022 the UK Government urged men to talk about mental health and engage with support networks, with suicide still reported to be the biggest cause of death in men under the age of 50 [18]. Previous evidence has shown that men are less likely than women to seek help for mental health problems and to engage with mental health services [19]. Future work should investigate the relative risk for individuals whose gender identity is different from their sex assigned at birth as previous literature has shown the risk is increased in this group [20].

Research investigating ethnic differences in the risk of suicide has produced inconsistent results [21]. Few studies from the UK have examined differences in the risk of suicide by ethnicity, although recent research found that White and Mixed ethnic groups had a higher suicide risk than Asian and Black ethnic groups [22]. Our results support these findings with suicide rate being highest in the White group compared to other ethnicities. Interestingly, previous work has highlighted complex interactions with other risk factors, for example, suicide risk is increased when individuals from ethnic minority groups live in areas with a low proportion of people from the same ethnic group.

Research has consistently found religion to be protective against suicide [23], [24], with social support shown to mediated the relationship between suicide ideation and attempts and religious affiliation. Research from Switzerland found suicide rates were highest among non-religious individuals [25]. In our study we find that the rates are lower among Muslim, Jewish, Sikh, Hindu and Christian groups compared to those who do not report having a religion. Conversely, for those who identify as Buddhist, or an alternative non-main religious group the rates are found to be higher than those who do not belong to a religious group. Several factors could be driving these differences in rates, such as differences in religious beliefs and behaviours between faiths, or unmeasurable differences between groups such as pre-existing predisposition to poor mental health. Reverse causality may also be at play, as it is possible that people in the UK are more likely to turn to Buddhism or other religions during times of distress. Religion is also a factor which is known to be changeable over time [27], with some groups being more likely to vary than others. Subsequent work should be conducted in the UK to investigate causal relationships between religious beliefs and suicide risk accounting for additional social and mental health factors.

Overall people who are unmarried experience a higher risk for suicide than married individuals [28]. Given the importance of episodic factors in relationship status, our finding of suicide rates being lower in individuals who were widowed compared to those who were single could be explained by survival bias. Previous work has shown that bereavement is a known risk factor for suicide, however the time between bereavement and suicide is important, with risk of suicide highest in the first week after death of a spouse [29]. Subsequent work is required in England and Wales to assess bereavement as a time-dependent covariate using population linked data.

### Strengths & limitations

The major strengths of this study include the use of a large, nationally representative cohort of adults who lived in England and Wales in 2011. The size of the cohort enabled us to explore differences in the risk of suicide across a wide range of socio-demographic factors for suicide, whilst also adjusting for other factors.

One of the main limitations of this work is we did not have any data on adverse life events, such as violence or abuse, bereavement, or job losses, which may be important factors affecting mental health and suicide risk [30]. In addition, we have no information on mental health conditions, which are likely to mediate the relationship between some sociodemographic characteristics and the risk of suicide. Previous research has shown that previous self-harm and suicide attempt are the most important predictors for subsequent suicide [31]. Future work should aim to link health data to administrative records to account for poor mental health prior to outcomes. In addition, our analysis identified sociodemographic risk factors at one point in time (2011 Census) and it would be beneficial to have longitudinal measurements of risk factors which are likely to change over time, such as day-to-day impairments, marital status, or socioeconomic position.

Furthermore, it is important to highlight that our population sample does not capture certain potentially vulnerable groups of people, such as those who migrated or did not link to patient demographic service (PDS), as all individuals in our study were enumerated in Census 2011.

## Conclusion

Our results show that suicide risk varies by sex and age, with males aged 40-to-50-years who are long term unemployed or have never worked, disabled or single having the highest rates. In women, we find a similar pattern, with the absolute rates being lower. Interestingly, the relative risk is greater for disabled people compared to non-disabled people for women, indicating that this group in particular should be targets of intervention. The current work provides novel population level insights into the groups with the highest rates and factors which are independently associated with suicide.

## Data sharing

In accordance with NHS Digital’s Information Governance requirements, the study data cannot be shared. Code used in this study is available on GitHub.

## Data Availability

In accordance with NHS Digitals Information Governance requirements the study data cannot be shared

## Supplementary tables

**Supplementary Table 1.** Sample Flow of population.

| **Stage** | **Count** |
| --- | --- |
| Sample which were valid Census respondents and could be linked to PDS | 50189388 |
| Sample were alive at end of study (31-10-2021), of had a data of death between 2011 Census day and EOS. | 50189220 |
| Sample who were between the ages of 18-74 years on 2011 Census day | 35136922 |
| Sample which had a positive time at risk | 35136916 |

**Supplementary Table 2.** Characteristics of study population and number of deaths.

| **Exposure** | **Group** | **Population size** | **Number of suicide deaths** | **Number of deaths from all other causes** |
| --- | --- | --- | --- | --- |
| Ethnicity | Arab | 97,134 | 26 | 1,893 |
| Ethnicity | Caribbean, African, Black British and other Black | 961,055 | 455 | 33,096 |
| Ethnicity | Chinese | 257,520 | 116 | 5,616 |
| Ethnicity | Indian | 919,616 | 561 | 31,007 |
| Ethnicity | Mixed/multiple ethnic groups | 457,277 | 592 | 13,475 |
| Ethnicity | Other ethnic group | 166,824 | 107 | 4,304 |
| Ethnicity | Pakistani and other Asian/Asian British | 1,258,557 | 470 | 32,465 |
| Ethnicity | White | 31,018,933 | 33,601 | 2,028,203 |
| Partnership status | In partnership | 18,001,454 | 12,466 | 1,197,359 |
| Partnership status | Partner deceased | 1,015,074 | 791 | 213,836 |
| Partnership status | Separated | 4,434,071 | 6,667 | 407,276 |
| Partnership status | Single | 11,686,317 | 16,004 | 331,588 |
| Religious affiliation | Buddhist | 166,336 | 230 | 5,574 |
| Religious affiliation | Christian | 21,240,630 | 18,811 | 1,556,782 |
| Religious affiliation | Hindu | 531,913 | 260 | 16,976 |
| Religious affiliation | Jewish | 149,274 | 125 | 7,548 |
| Religious affiliation | Muslim | 1,381,452 | 411 | 36,028 |
| Religious affiliation | No religion | 8,962,297 | 12,673 | 345,882 |
| Religious affiliation | Not stated | 2,251,635 | 2,812 | 161,440 |
| Religious affiliation | Other | 183,955 | 332 | 9,838 |
| Religious affiliation | Sikh | 269,424 | 274 | 9,991 |
| Day-to-day impairments | Day to day activities limited a lot/a little | 5,818,942 | 10,880 | 1,136,315 |
| Day-to-day impairments | Day to day activities not limited | 29,317,974 | 25,048 | 1,013,744 |
| Age group | 18-29 | 7,392,373 | 6,627 | 38,232 |
| Age group | 30-39 | 6,469,695 | 7,152 | 72,528 |
| Age group | 40-49 | 7,515,790 | 9,914 | 198,543 |
| Age group | 50-59 | 6,358,710 | 6,874 | 412,186 |
| Age group | 60-69 | 5,734,463 | 4,248 | 909,560 |
| Age group | 70-79 | 1,665,885 | 1,113 | 519,010 |
| Region | East of England | 3,701,430 | 3,609 | 215,978 |
| Region | London | 4,828,097 | 3,824 | 211,299 |
| Region | North East | 1,676,937 | 1,981 | 123,536 |
| Region | North Midlands | 2,912,812 | 2,868 | 187,354 |
| Region | North West | 4,466,715 | 5,043 | 316,806 |
| Region | South East | 5,437,626 | 5,452 | 303,875 |
| Region | South West | 3,364,081 | 3,705 | 205,442 |
| Region | Wales | 1,919,728 | 2,204 | 134,751 |
| Region | West Midlands | 3,499,287 | 3,577 | 228,900 |
| Region | Yorkshire and the Humber | 3,330,203 | 3,665 | 222,118 |
| Armed forces member | Dependent (spouse, same-sex civil partner,<br>partner, child<br>or stepchild) of<br>member of<br>armed forces | 61,084 | 28 | 589 |
| Armed forces<br>member | Member of<br>armed forces | 50,352 | 56 | 748 |
| Armed forces<br>member | Not member or<br>dependent of<br>armed forces | 35,025,480 | 35,844 | 2,148,722 |
| National<br>Statistics Socio-economic<br>classification | Class 1.1 | 1,191,885 | 833 | 49,203 |
| National<br>Statistics Socio-economic<br>classification | Class 1.2 | 3,720,296 | 2,938 | 130,840 |
| National<br>Statistics Socio-economic<br>classification | Class 2 | 8,169,305 | 7,049 | 363,646 |
| National<br>Statistics Socio-economic<br>classification | Class 3 | 3,635,277 | 3,448 | 212,166 |
| National<br>Statistics Socio-economic<br>classification | Class 4 | 4,558,275 | 4,922 | 270,258 |
| National<br>Statistics Socio-economic<br>classification | Class 5 | 3,108,307 | 3,200 | 214,207 |
| National<br>Statistics Socio-economic<br>classification | Class 6 | 4,452,001 | 5,238 | 353,227 |
| National<br>Statistics Socio-economic<br>classification | Class 7 | 4,235,312 | 5,271 | 401,413 |
| National<br>Statistics Socio-economic<br>classification | Class 8 | 1,103,609 | 1,928 | 99,379 |
| National<br>Statistics Socio-economic<br>classification | Students | 567,382 | 421 | 6,013 |
| National<br>Statistics Socio-economic<br>classification | Not classifiable | 395,267 | 680 | 49,707 |
| Sex | Female | 18,173,538 | 9,393 | 926,684 |
| Sex | Male | 16,963,378 | 26,535 | 1,223,375 |

**Supplementary table 3.**
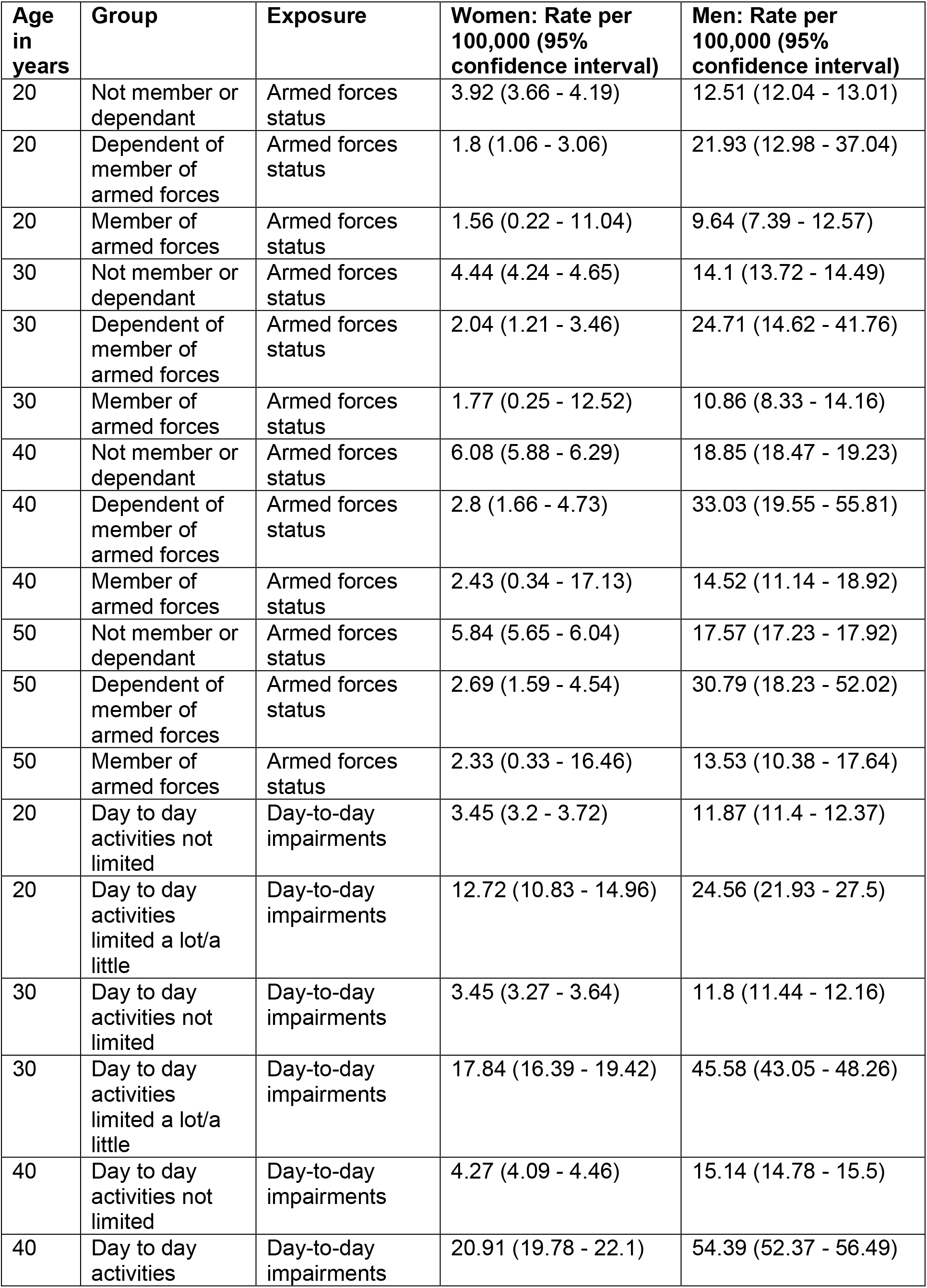
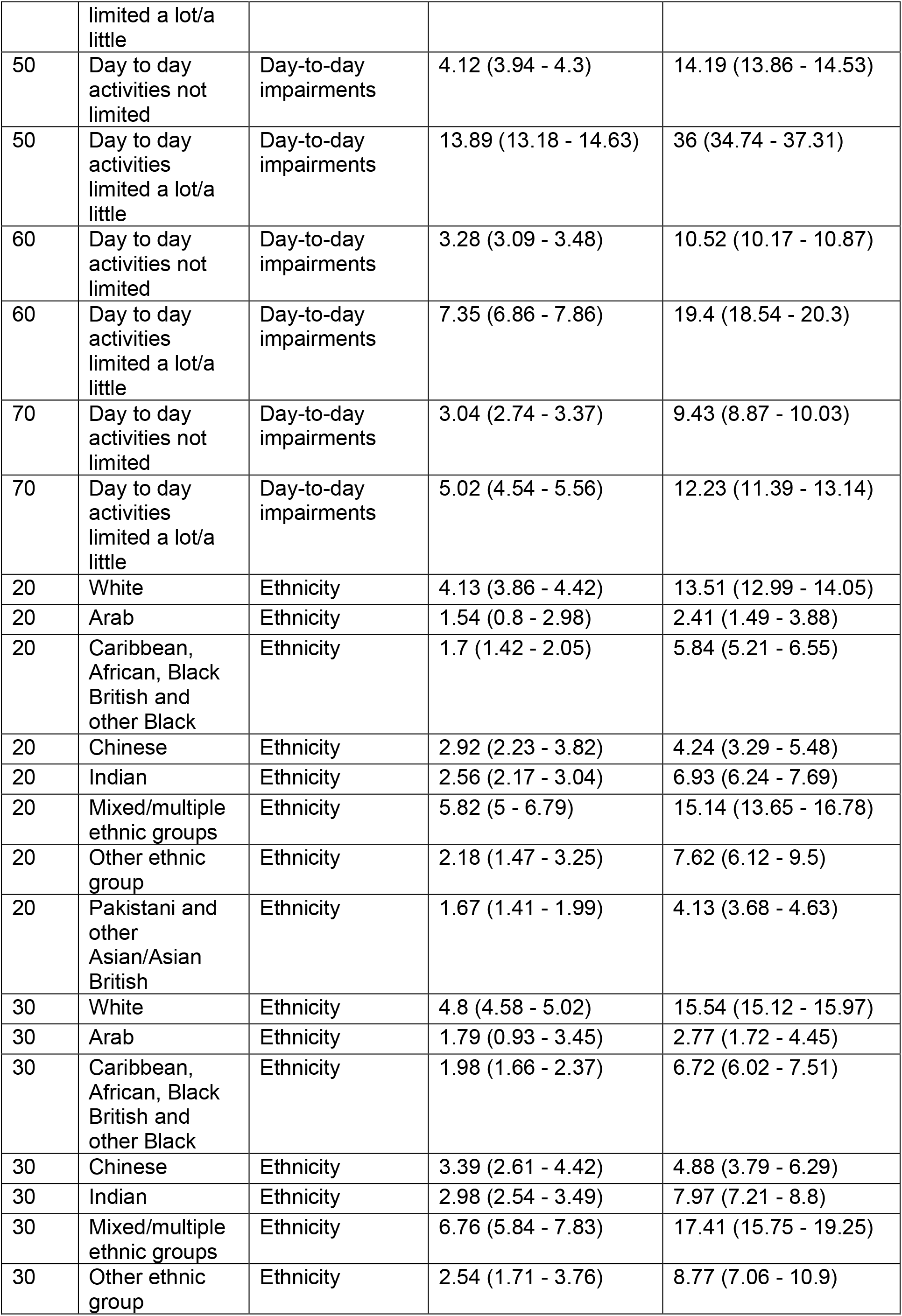
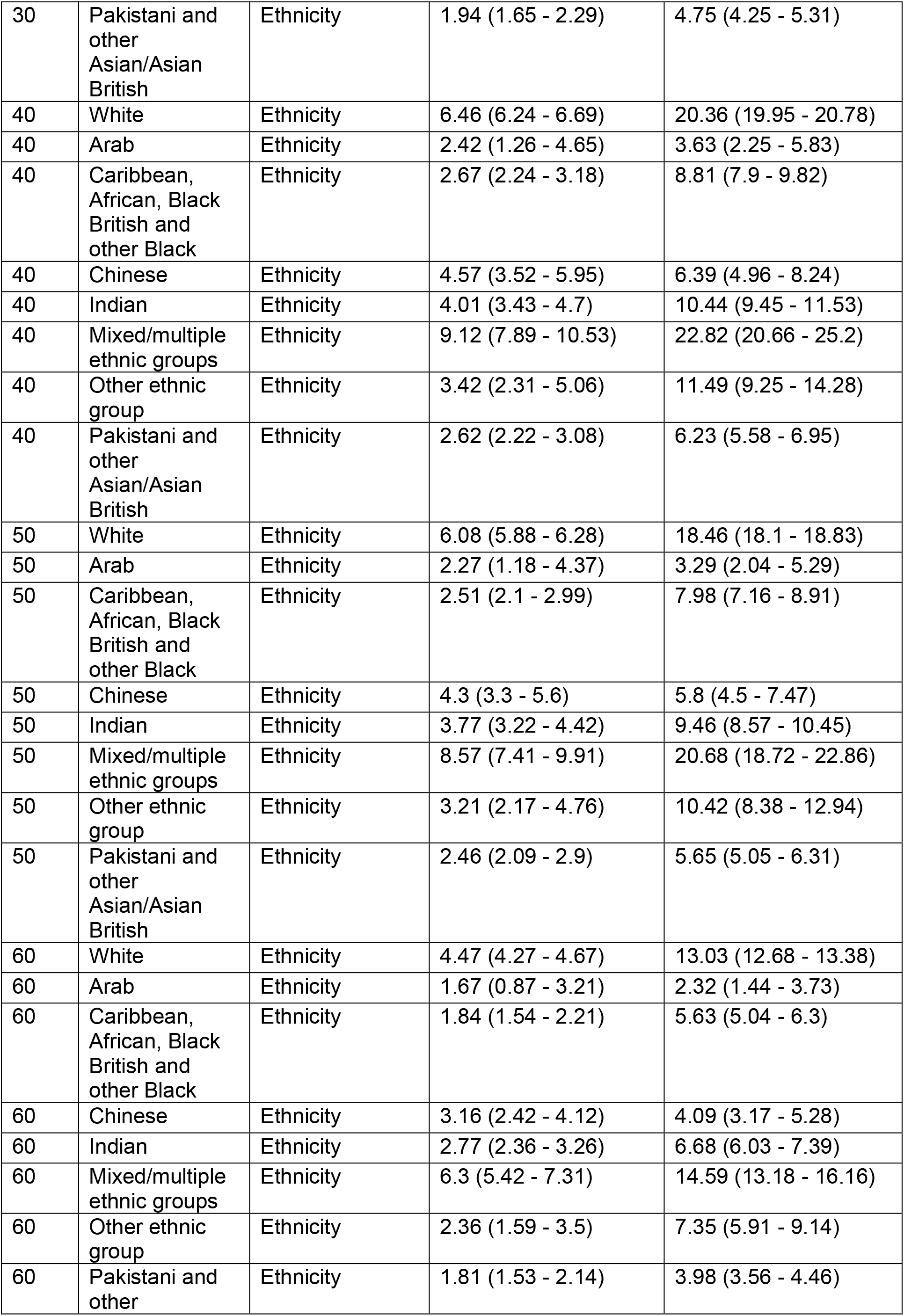
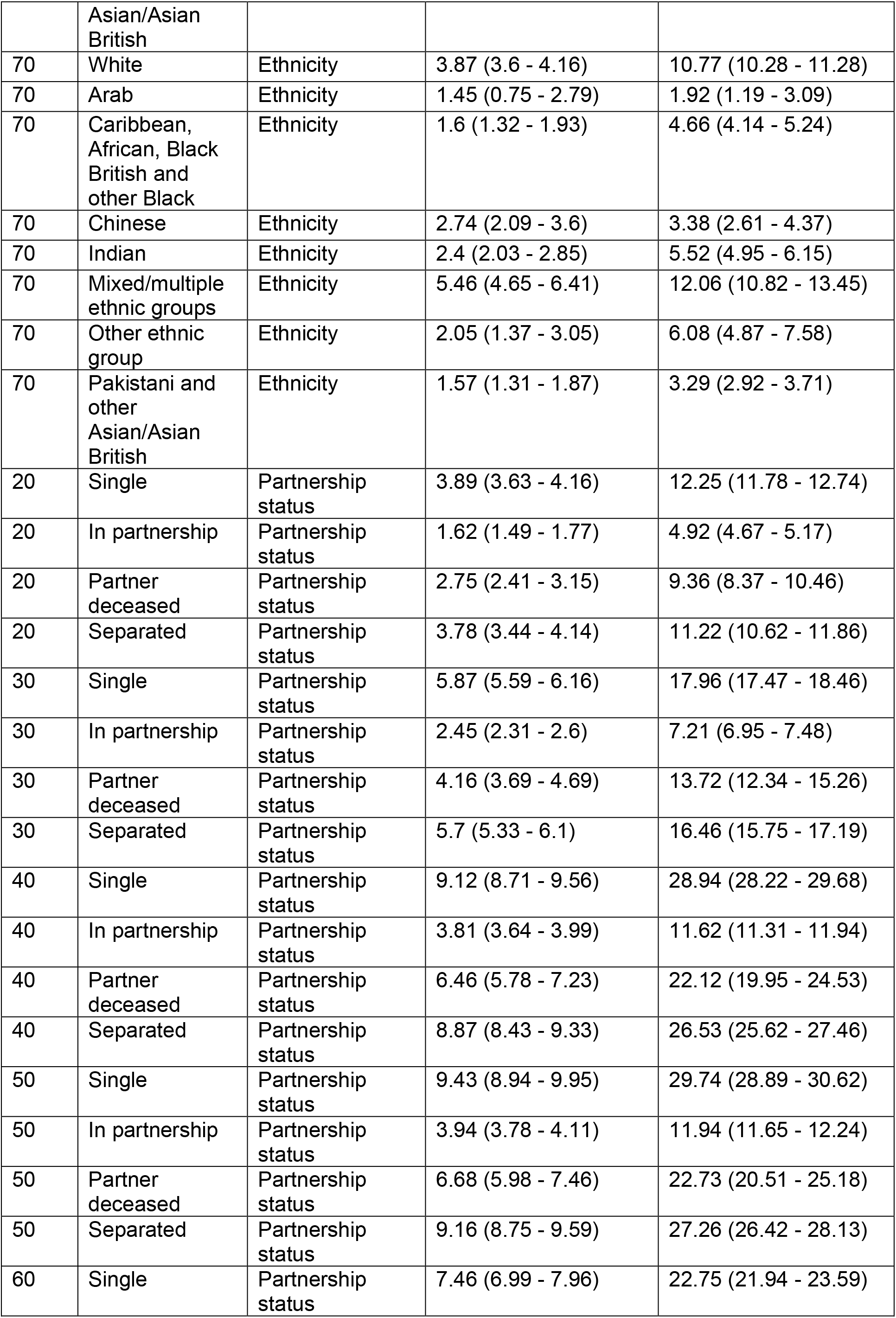
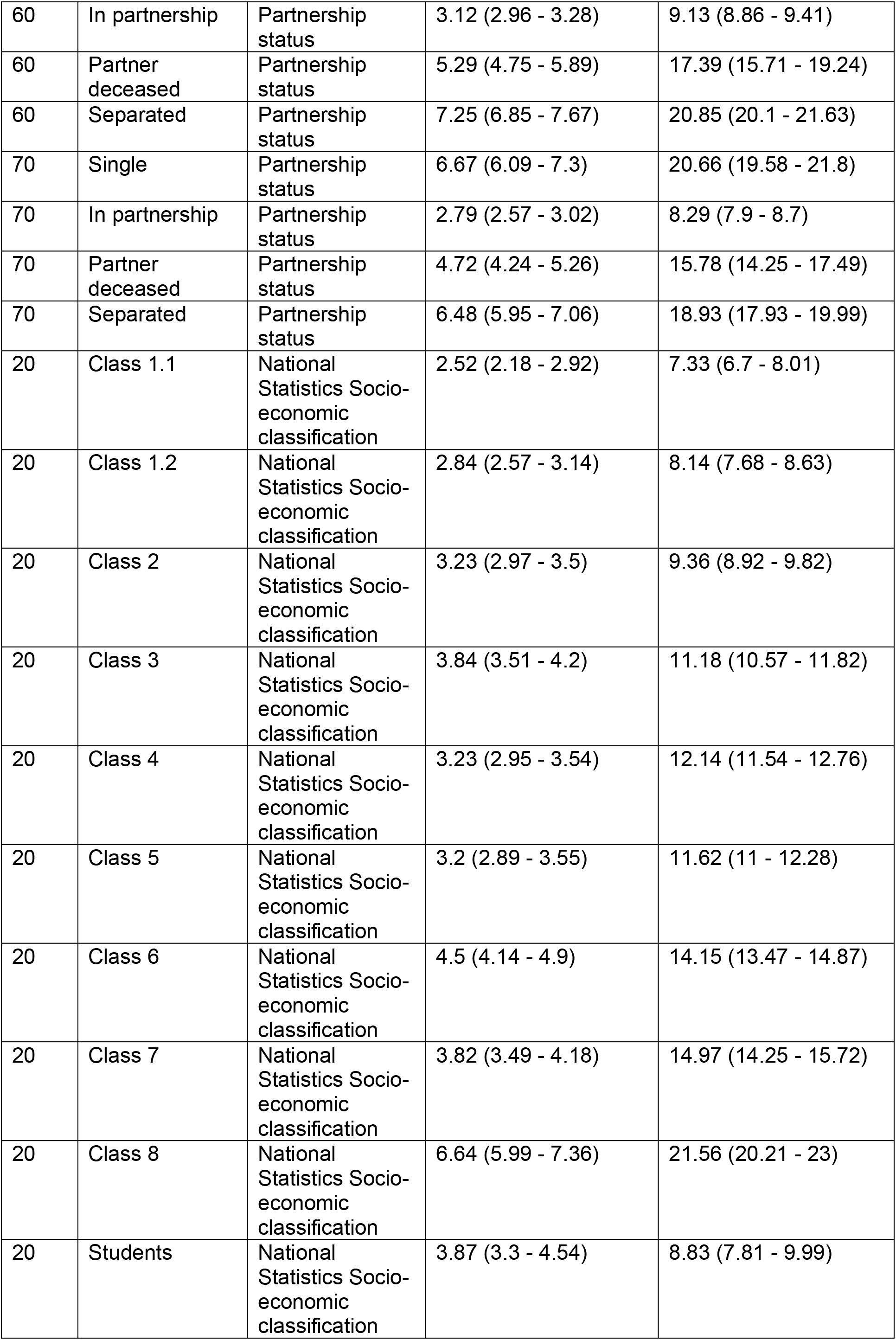
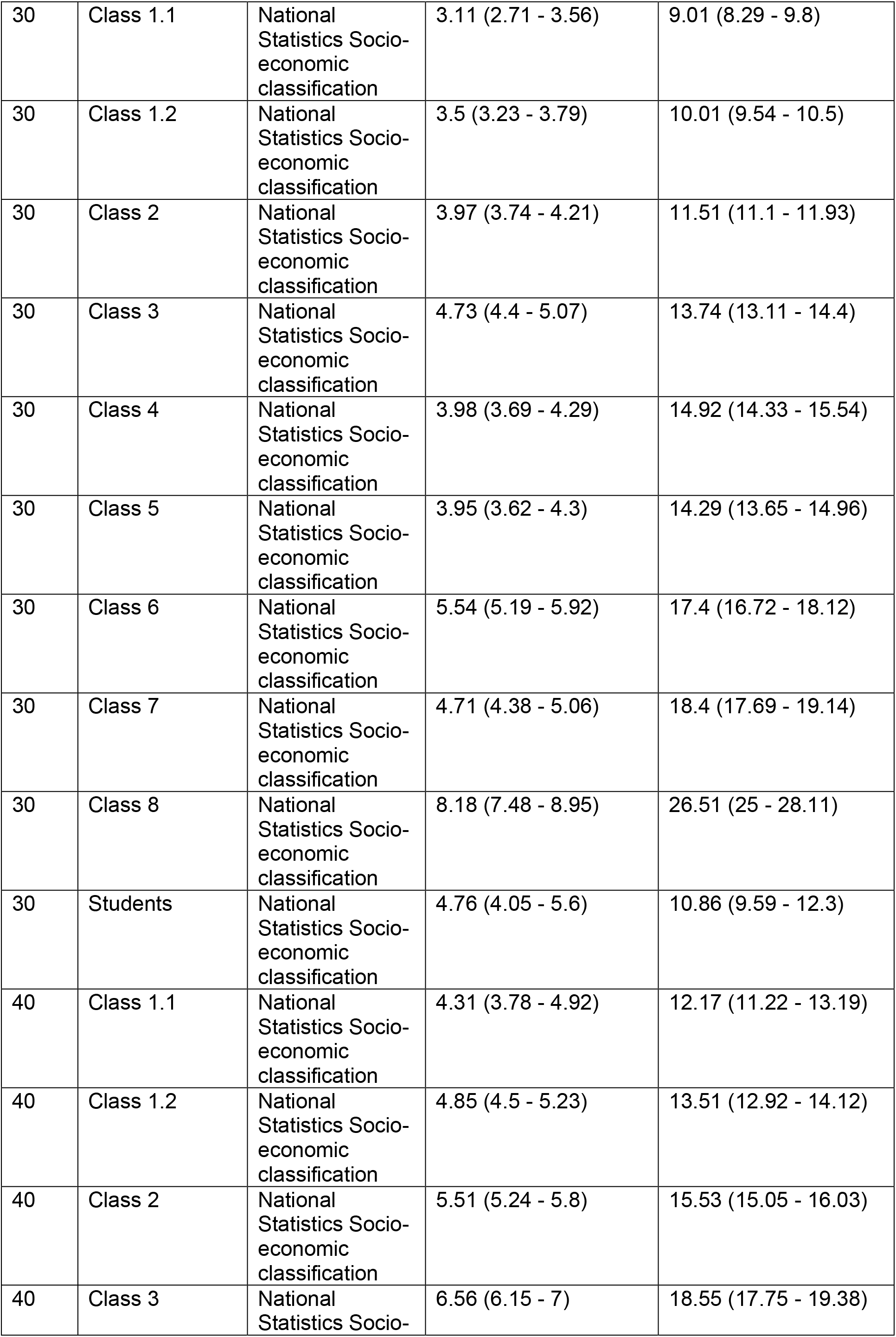
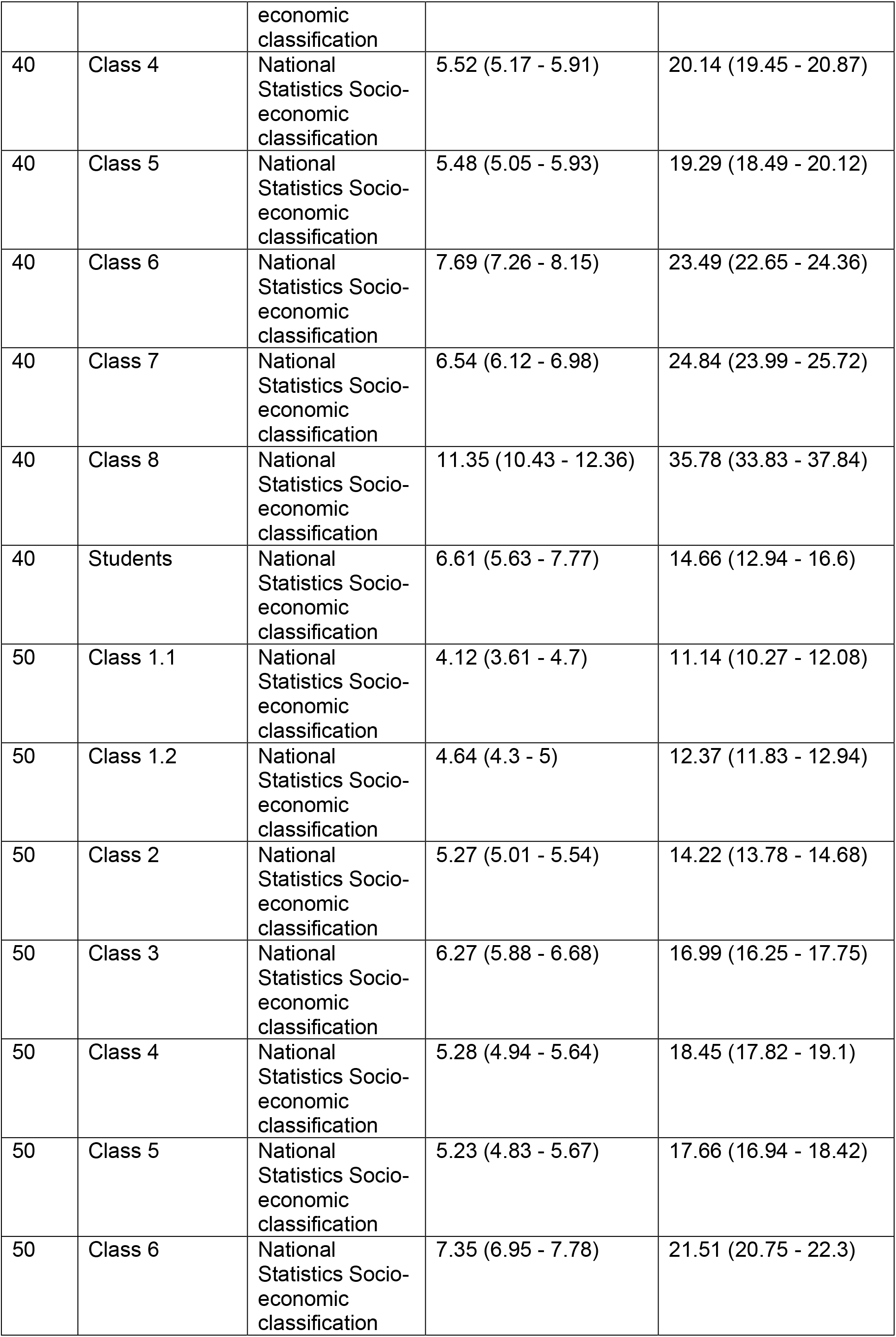
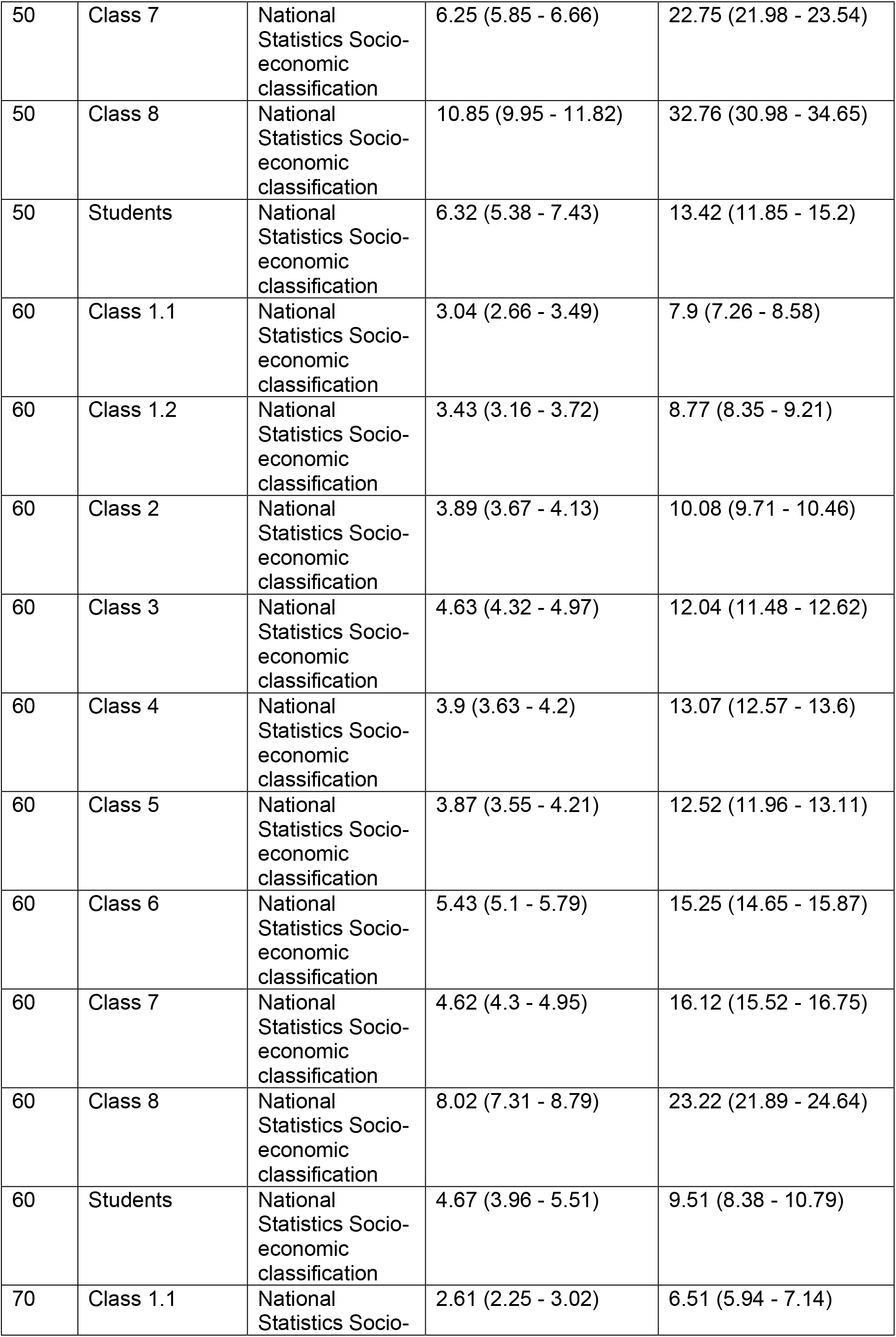
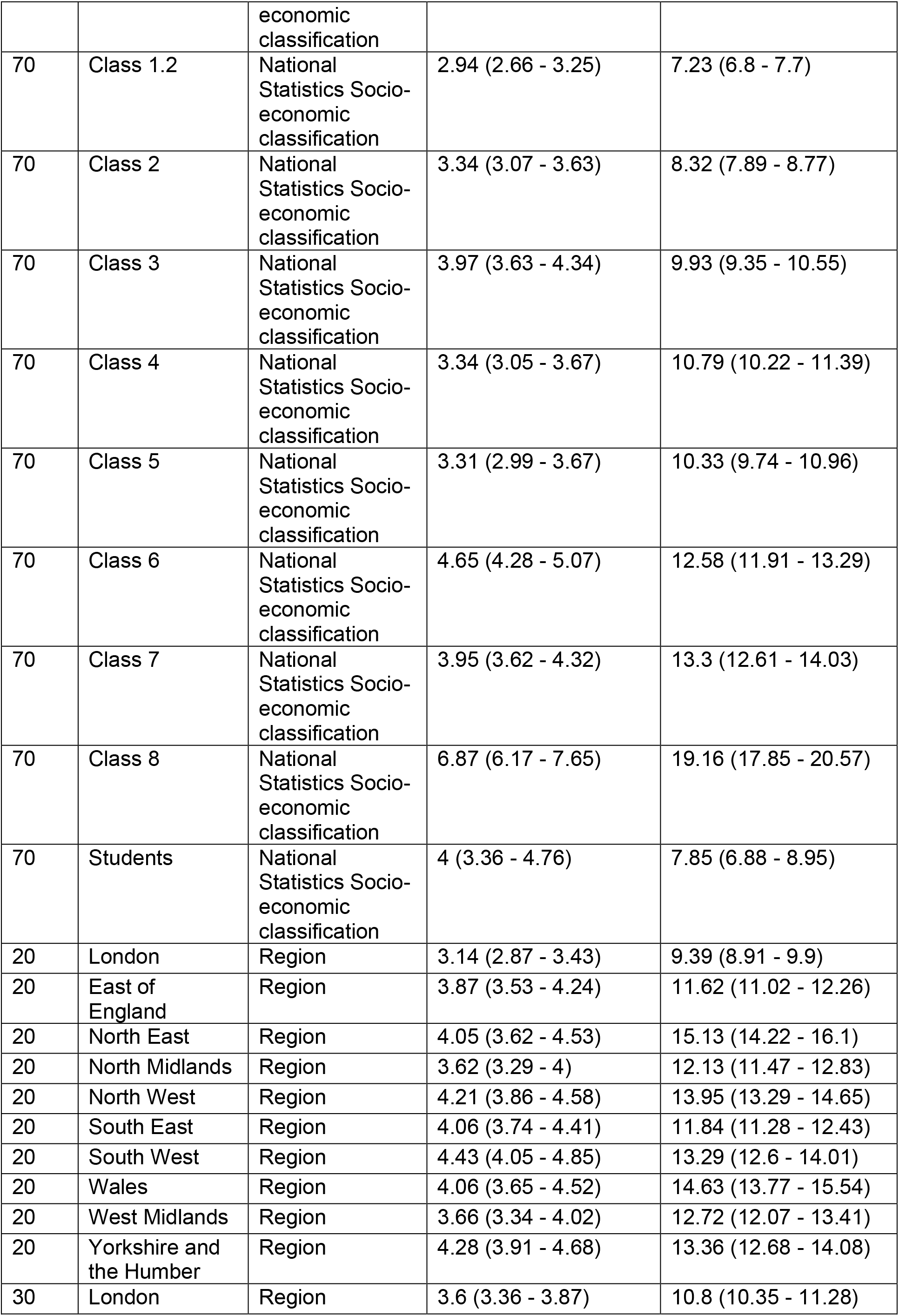
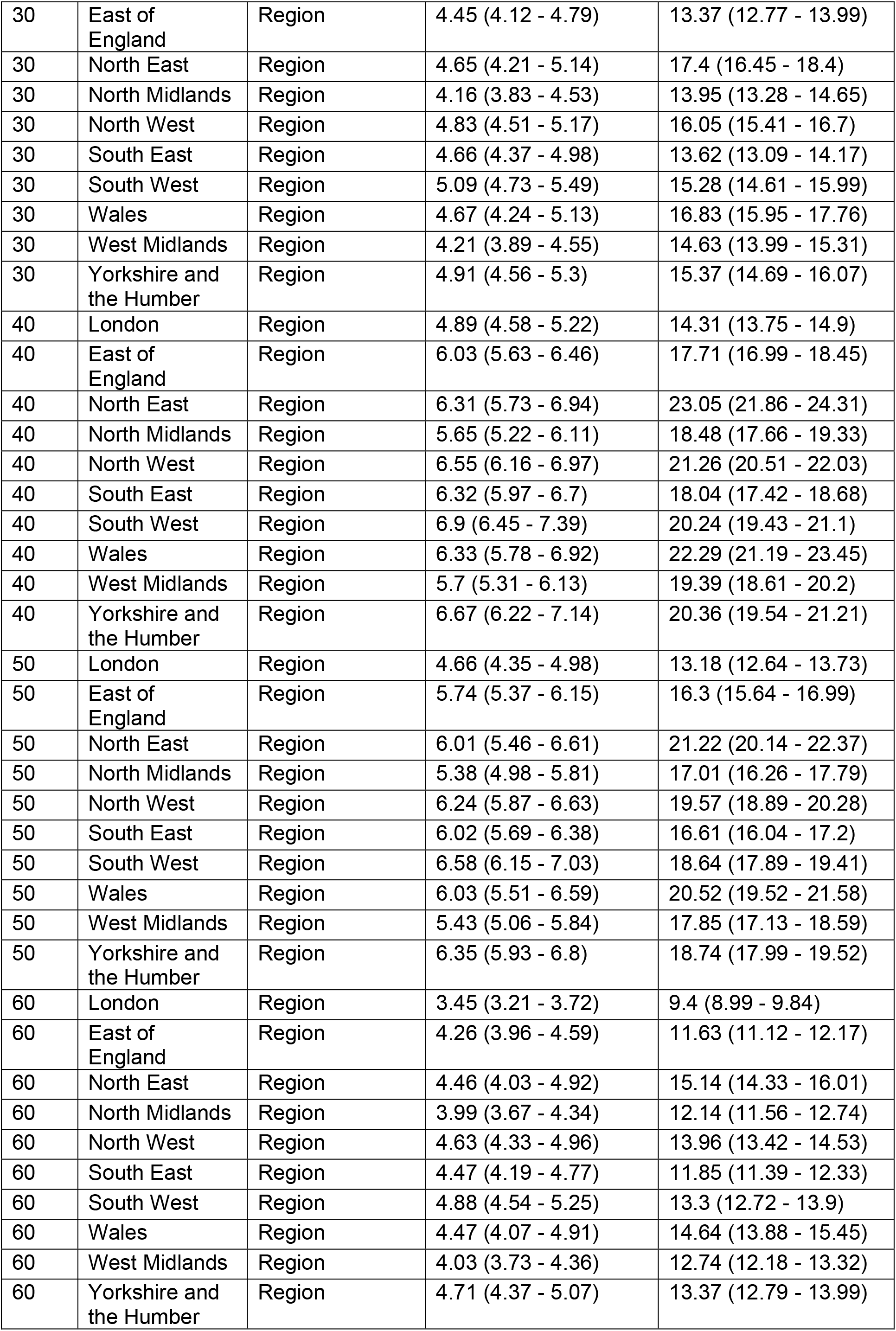
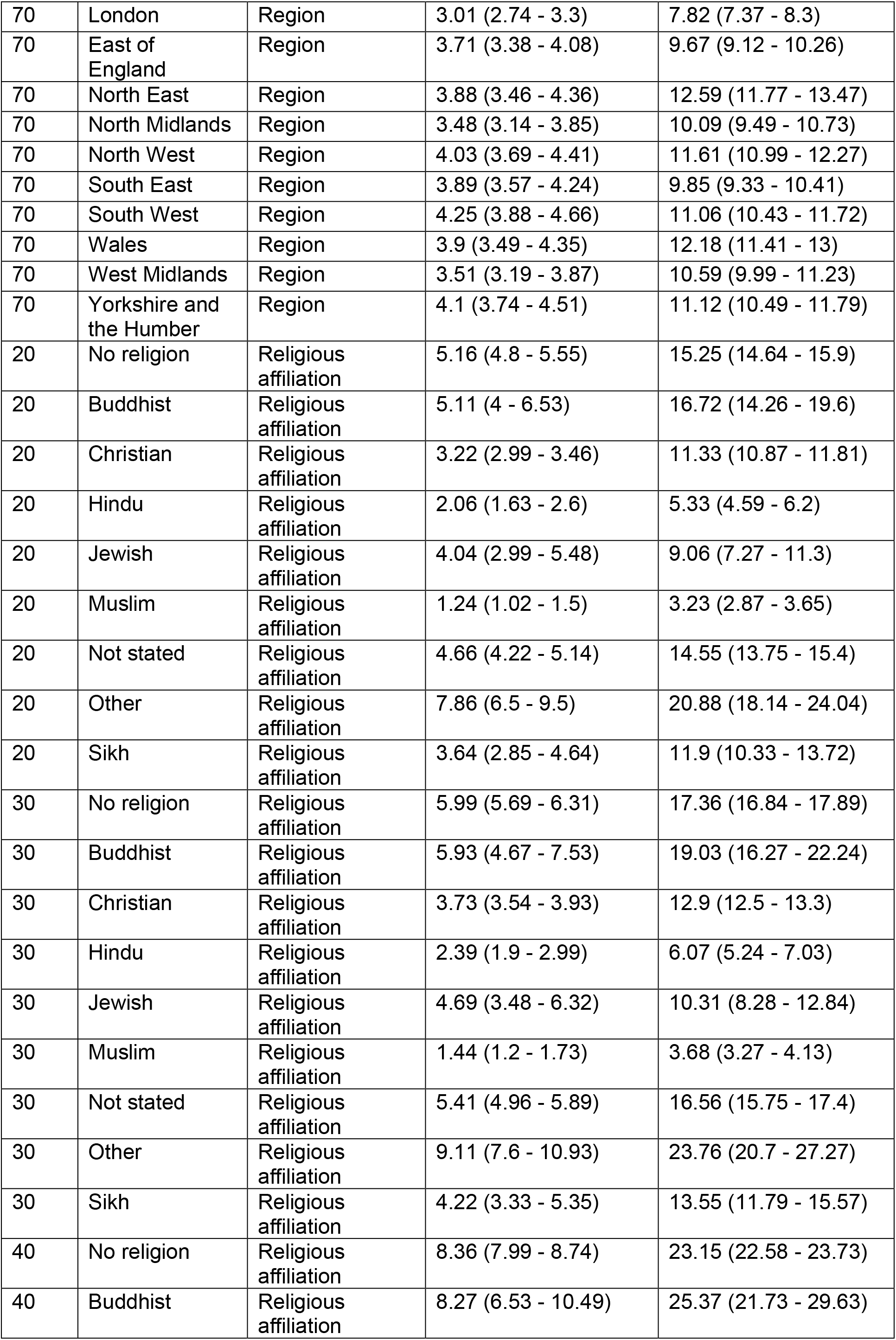
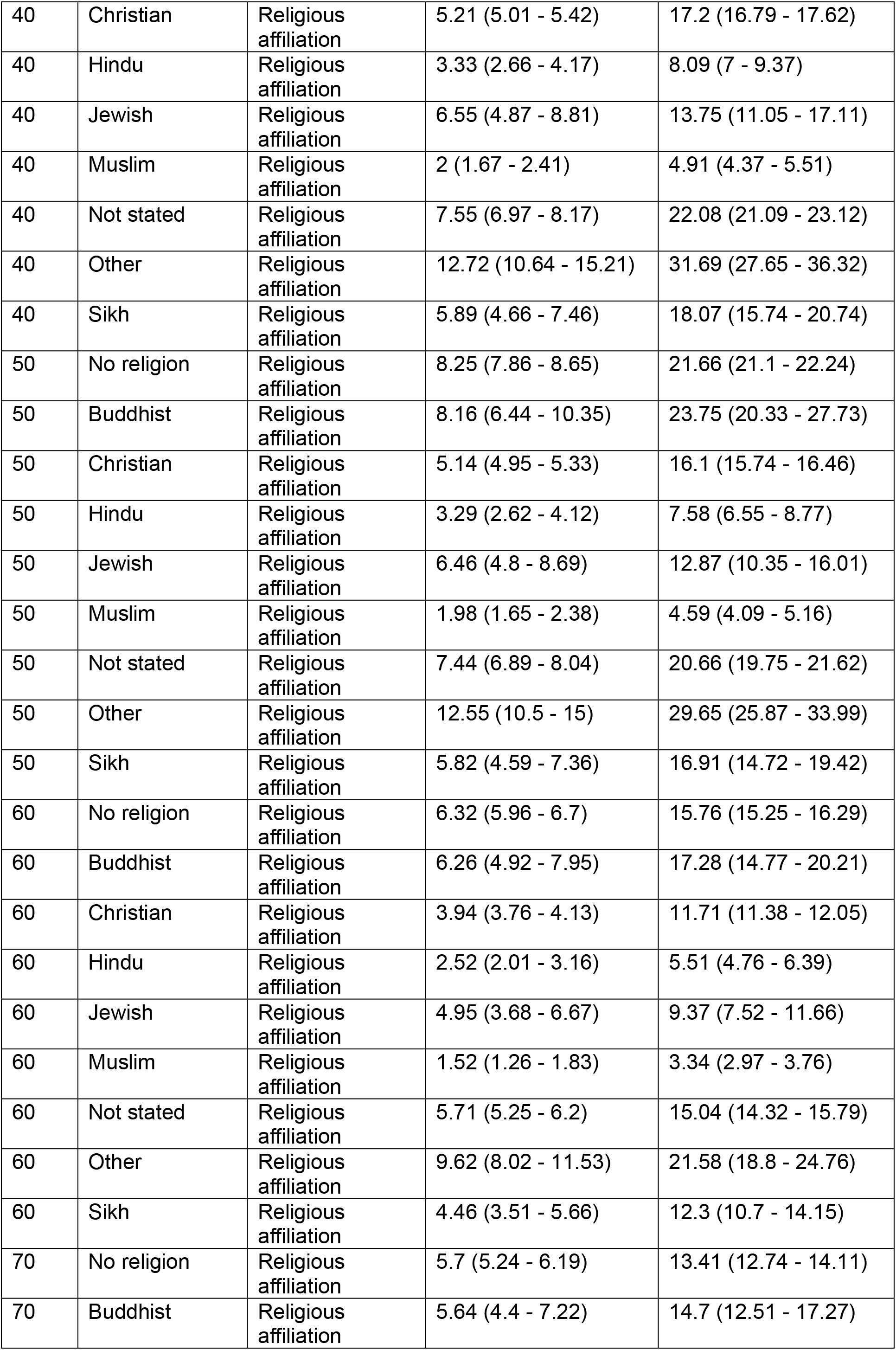
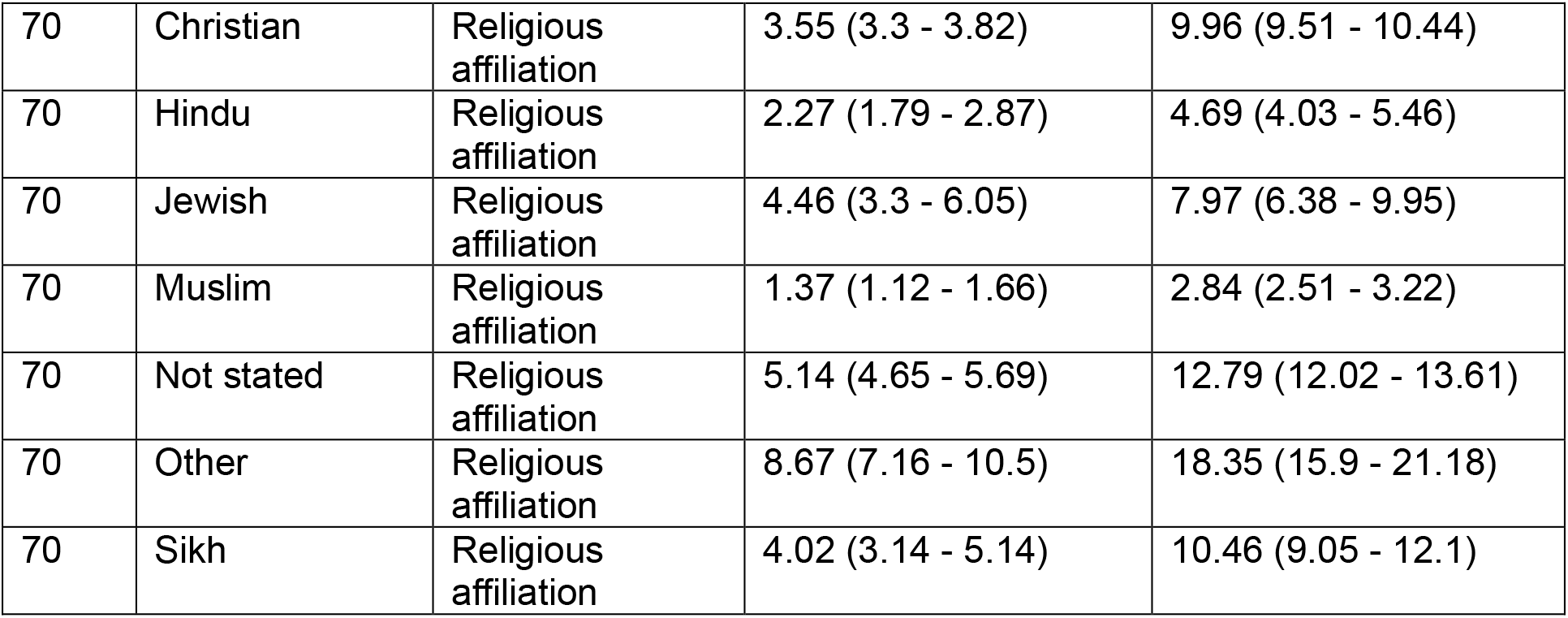
Estimated rates of suicide by sociodemographic characteristics for average age.

